# The Gene Curation Coalition: A global effort to harmonize gene-disease evidence resources

**DOI:** 10.1101/2022.01.03.21268593

**Authors:** Marina T. DiStefano, Scott Goehringer, Lawrence Babb, Fowzan S. Alkuraya, Joanna Amberger, Mutaz Amin, Christina Austin-Tse, Marie Balzotti, Jonathan S. Berg, Ewan Birney, Carol Bocchini, Elspeth A. Bruford, Alison J. Coffey, Heather Collins, Fiona Cunningham, Louise C. Daugherty, Yaron Einhorn, Helen V. Firth, David R. Fitzpatrick, Rebecca E. Foulger, Jennifer Goldstein, Ada Hamosh, Matthew R. Hurles, Sarah E. Leigh, Ivone US. Leong, Sateesh Maddirevula, Christa L. Martin, Ellen M. McDonagh, Annie Olry, Arina Puzriakova, Kelly Radtke, Erin M. Ramos, Ana Rath, Erin Rooney Riggs, Angharad M. Roberts, Charlotte Rodwell, Catherine Snow, Zornitza Stark, Jackie Tahiliani, Susan Tweedie, James S. Ware, Phillip Weller, Eleanor Williams, Caroline F. Wright, T Michael. Yates, Heidi L. Rehm

**Affiliations:** Geisinger Health System, Danville, PA, USA; Program in Medical and Population Genetics, Broad Institute of MIT and Harvard, Cambridge, MA, USA; Department of Translational Genomics, Center for Genomic Medicine, King Faisal Specialist Hospital and Research Center, Riyadh, 11211, Saudi Arabia; Department of Genetic Medicine, Online Mendelian Inheritance in Man (OMIM), Johns Hopkins University School of Medicine, Baltimore, MD, 21287-4922, USA; Inserm, US14 - Orphanet, France; Department of Pathology, Massachusetts General Hospital, Boston, MA, USA; MGB Laboratory for Molecular Medicine, Cambridge, MA, USA; Myriad Women’s Health, San Francisco, CA, USA; Department of Genetics, University of North Carolina, Chapel Hill, NC, USA; European Molecular Biology Laboratory, European Bioinformatics Institute, Wellcome Genome Campus, Hinxton, CB10 1SD, UK; HUGO Gene Nomenclature Committee (HGNC), European Molecular Biology Laboratory, European Bioinformatics Institute, Wellcome Genome Campus, Hinxton, CB10 1SD, UK; Department of Haematology, University of Cambridge School of Clinical Medicine, Cambridge, CB2 0PT, UK; Illumina Clinical Services Laboratory, Illumina Inc., 5200 Illumina Way, San Diego, CA, 92122, USA; National Library of Medicine, Bethesda, MD, USA; ICF, 9300 Lee Highway, Fairfax, VA, 22031, USA; Genome Interpretation, Genome Assembly and Annotation (GAA), European Molecular Biology Laboratory, European Bioinformatics Institute,, Wellcome Genome Campus, Hinxton,, Cambridge, CB10 1SD, UK; Genomics England, Queen Mary University of London, Dawson Hall, Charterhouse Square, London, EC1M 6BQ, UK; Healx Ltd., Charter House, 66-68 Hills Rd, Cambridge, CB2 1LA, UK; Franklin by Genoox, Palo Alto, CA, USA; Department of Genetics, Addenbrooke’s Hospital, Cambridge, UK; MRC Human Genetics Unit, MRC IGMM, The University of Edinburgh, Edinburgh, UK; SciBite Limited, BioData Innovation Centre, Wellcome Genome Campus, Hinxton, CB10 1DR, UK; Wellcome Sanger Institute, Hinxton, UK; Open Targets, EMBL-EBI, Wellcome Genome Campus, Hinxton, CB10 1DR, UK; AmbryGenetics, Aliso Viejo, CA, USA; National Human Genome Research Institute, National Institutes of Health, USA; Autism & Developmental Medicine Institute, Geisinger Health System, Danville, PA, USA; National Heart & Lung Institute & MRC London Institute of Medical Sciences, Imperial College London, London, UK; Great Ormond Street Hospital, London, UK; Australian Genomics, Melbourne, Australia; Invitae, San Francisco, CA, USA; Royal Brompton & Harefield Hospitals, Guy’s and St. Thomas’ NHS Foundation Trust, London, UK; Institute of Biomedical and Clinical Science, University of Exeter Medical School, Royal Devon & Exeter Hospital, Exeter, EX2 5DW, UK; Center for Genomic Medicine, Massachusetts General Hospital, Boston, MA, USA

## Abstract

**PURPOSE:** Several groups and resources provide information that pertains to the validity of gene-disease relationships used in genomic medicine and research; however, universal standards and terminologies to define the evidence base for the role of a gene in disease, and a single harmonized resource were lacking. To tackle this issue, the Gene Curation Coalition (GenCC) was formed.

**METHODS:** The GenCC drafted harmonized definitions for differing levels of gene-disease validity based on existing resources, and performed a modified Delphi survey with three rounds to narrow the list of terms. The GenCC also developed a unified database to display curated gene-disease validity assertions from its members.

**RESULTS:** Based on 241 survey responses from the genetics community, a consensus term set was chosen for grading gene-disease validity and database submissions. As of December 2021, the database contains 15,241 gene-disease assertions on 4,569 unique genes from 12 submitters. When comparing submissions to the database from distinct sources, conflicts in assertions of gene-disease validity ranged from 5.3% to 13.4%.

**CONCLUSION:** Terminology standardization, sharing of gene-disease validity classifications, and resolution of curation conflicts will facilitate collaborations across international curation efforts and in turn, improve consistency in genetic testing and variant interpretation.

## Introduction

### Rationale for formation of GenCC

With the decreasing cost of sequencing, exome and genome analysis has become more common for many clinical indications, necessitating gene-level knowledge on a larger list of genes with an increasing scientific curation burden. The assessment of the evidence that variants in a gene are linked to a particular monogenic disease is critical for variant interpretation and determining the content for clinical gene panel tests. Unless a gene is convincingly linked to disease, the pathogenicity of a variant cannot be interpreted.^1-3^ Thus, curation of gene-disease validity is a fundamental prerequisite for classifying variants identified in a variety of contexts such as diagnostic genetic testing and disease risk screening as well as determining the genetic basis for all human diseases. Several groups and resources provide information that pertains to the validity of gene-disease relationships; however, the standards and terminologies to define the evidence base for a gene’s role in disease were not harmonized. To tackle this issue, the Gene Curation Coalition (GenCC, www.thegencc.org) was formed including organizations that provide online gene-level resources as well as diagnostic laboratories that have committed to sharing their internally curated gene-level knowledge. Together, this group is working to standardize approaches to ensure gene-level resources are interoperable and concordant, allowing groups to work together most effectively and providing consistent and useful resources for the community. Harmonization efforts of the GenCC began with clarifying the curation processes used by member groups, followed by the development of consistent terminology for clinical validity assessment, inheritance, allelic requirement, and mechanism of disease.

Work with sequence and copy number variants has demonstrated that knowledge sharing is critical^4,5^ and can help resolve classification discrepancies,^6-9^ particularly through ClinVar,^10^ the community’s primary database for sharing variant-level assertions. In addition, UK and Australian groups have demonstrated the value of collaborating on gene-disease assertions to define gene panels for clinical testing.^11^ Here we demonstrate the launch of an international database of gene-disease validity assertions to further facilitate data sharing and resolve discrepancies in assertions of gene-disease validity.

### GenCC member groups

The GenCC consists of databases that provide public gene level resources as well as diagnostic laboratories that have committed to sharing their internally curated gene-disease validity knowledge. The current members of the GenCC are as follows: Ambry Genetics, Clinical Genome Resource (ClinGen), DECIPHER, Franklin by Genoox, Genomics England PanelApp, HGNC (HUGO Gene Nomenclature Committee), Illumina Inc., Invitae, King Faisal Specialist Hospital and Research Center, Mass General Brigham Laboratory for Molecular Medicine, Myriad Women’s Health, OMIM (Online Mendelian Inheritance in Man), Orphanet, PanelApp Australia, and the Gene2Phenotype Database of TGMI (Transforming Genomic Medicine Initiative). Descriptions of each group’s curation efforts and steering committee membership can be found on the GenCC website (https://thegencc.org/members.html).

## Materials and Methods

### GenCC Logistics

The GenCC was formed in February 2018 during a joint meeting of the Transforming Genetic Medicine Initiative and ClinGen at the Wellcome Trust, London, UK. After this first in-person meeting, two more in-person meetings took place at the Curating the Clinical Genome Conferences in 2018 in Hinxton, UK, and in 2019 in Washington DC, USA. In addition, the group meets monthly via web conferencing.

### Harmonizing Gene-Disease Validity definitions

Each GenCC member group presented an overview of their resource including: audience, purpose, and curation content. All GenCC members worked together to generate a list of harmonized definitions of gene-disease validity (Table S1) to use in a modified Delphi survey for standardizing these terms for display in the GenCC database (Figure 1). While many member groups had their own separate gene-disease validity definitions in their own resources, the common definitions, which included concrete examples of evidence levels, were useful for developing consensus terminologies (Table S1). A final version of these definitions that includes the harmonized validity terms is also found on the GenCC website (https://thegencc.org/faq.html#validity-termsdelphi-survey).

**Figure 1:**
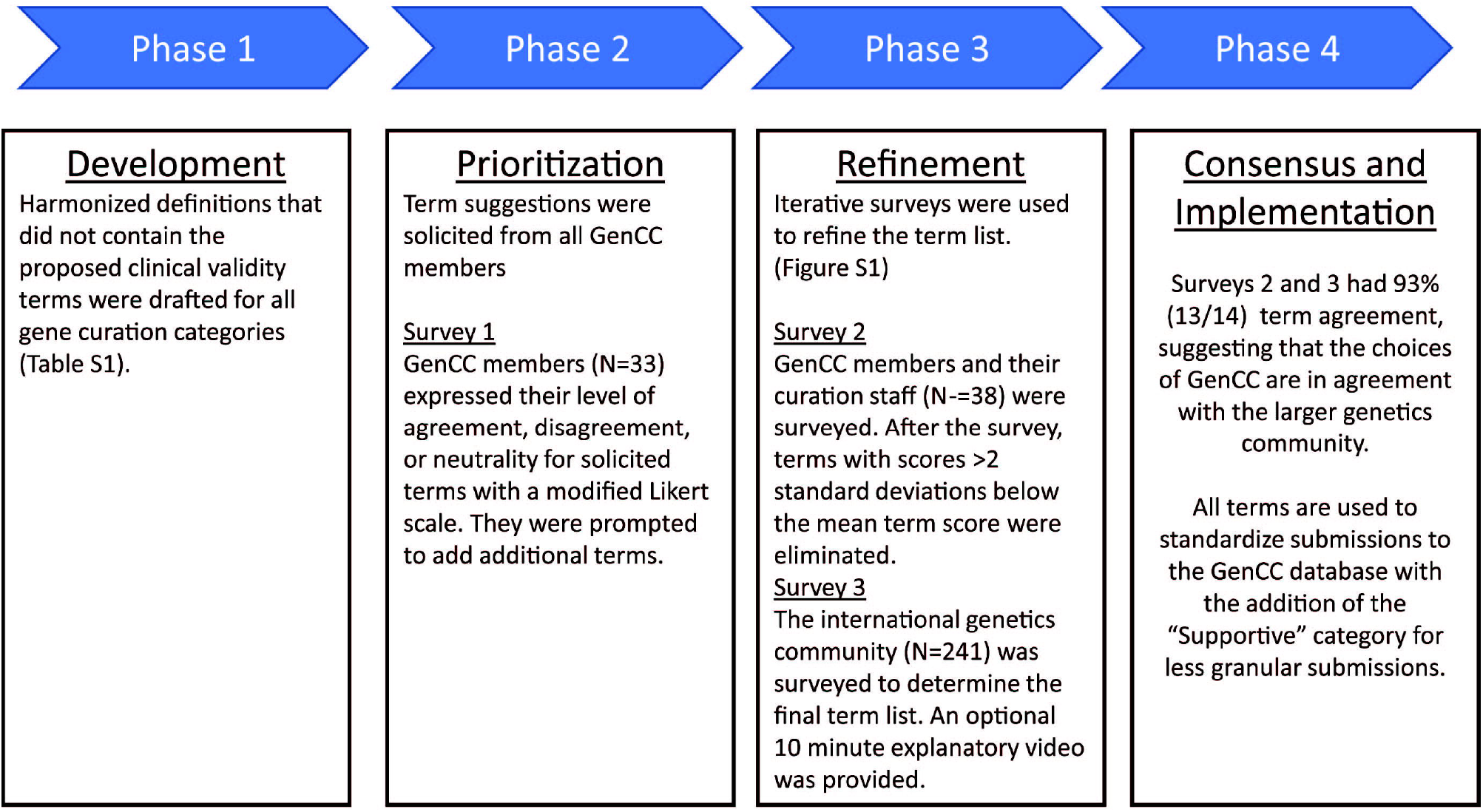
Phases of the modified Delphi survey process for GenCC gene-disease validity terminology.

### Evidence v Likelihood terms

While drafting common definitions for gene-disease validity terms, the GenCC member groups realized that they referred to gene disease validity with two different types of terms (Table S2). One range of terms described the evidence present for a gene-disease pair (denoted “evidence terms” in the Delphi survey), examples of which include *limited, moderate*, and *strong*. The second set of terms used by member groups described the confidence or likelihood that a gene was related to disease (denoted “likelihood terms” in the Delphi survey), examples of which include *confirmed, possible, probable*, and *likely*. Both term sets were considered and separated in the Delphi survey.

### Generating a Delphi Survey

A first draft used a modified Delphi approach similar to the survey used to choose terms for clinical pharmacogenetic test results offered by the Clinical Pharmacogenetics Implementation Consortium (CPIC) (Figure 1).^12^ Participants were first asked about their demographic information; the demographic questions were based on those used in OMIM user surveys and the American Society of Human Genetics (ASHG) membership form. Participants were then asked to choose terms for differing levels of gene-disease validity, and finally rank previously generated term lists. Questions from the three iterations of the survey are present in the supplement. In round 1, the survey was taken by GenCC members and additional gene-disease validity terms were solicited from survey takers. In round 2, the survey was distributed to the extended membership of the GenCC groups. It was accompanied by an optional ∼8 minute video (https://vimeo.com/306463165) explaining the GenCC and rationale behind the survey. At the conclusion of the survey, those terms that were most unpopular (N=16) were eliminated. This was determined by assigning a numerical value to each Likert response: “strongly agree” (2), “agree” (1), neutral (0), disagree (−1), strongly disagree (−2). For each question all answers were summed, and a mean and standard deviation were calculated. All terms with a score greater than two standard deviations below the mean were eliminated if the question contained more than two term options (Figure S1). In round 3, the survey content was finalized and sent with the optional video to the genetics community, including 10 groups: the European Society of Human Genetics (ESHG), ASHG, the Canadian College of Medical Genetics (CCMG), ClinGen, the Association for Clinical Genomic Science (ACGS), the British Society for Genetic Medicine (BSGM), the Association for Molecular Pathology (AMP), the Association of Genetic Nurses and Counsellors (AGNC), Australian Genomics, and users of PanelApp, as well as posting on Twitter.

### Database

The GenCC database (https://search.thegencc.org/) is built using a MySQL backend server and was launched in December 2020. All submitters to the database are GenCC members. New members are welcome and are added to GenCC after demonstrating work consistent with evidence-based evaluation of gene-disease validity. Assertions are mapped to the standardized terms chosen by the modified Delphi survey. All submissions include a validity claim on a gene, disease, and a mode of inheritance (MOI). More information and database screenshots are provided in the results section. GenCC data is freely and openly available via download files from the GenCC website (https://search.thegencc.org). An API interface providing additional flexibility and expanded access to the GenCC database is planned for later in 2022.

The GenCC website also provides an easy to use query and display interface for interactive usage, designed for researchers and clinicians to quickly access information regarding genes of interest. The underlying database supporting the GenCC website contains all submitted assertion data and is optimized for rapid query results. To ensure the integrity and accurate representation of all data, submitted assertions are held to strict formatting and evidence requirements and reviewed by the GenCC staff prior to publishing on the website.

### Submission Process

The primary attributes of an assertion are a gene, disease, mode of inheritance, classification, assertion date, and submitter. A formatted spreadsheet, conceptually similar to a ClinVar submission spreadsheet (https://ftp.ncbi.nlm.nih.gov/pub/clinvar/submission_templates/) is used for GenCC database submission. The most recent version of this sheet can be found on the website (https://thegencc.org/submission-directions). In this spreadsheet, users include standard ontology or identifiers to input each gene (HGNC ID), disease (OMIM Phenotype MIM number, ORPHAcode, or Monarch Disease Ontology (MONDO) identifier), mode of inheritance (human phenotype ontology (HPO) term), validity assertion (standardized terms from the modified Delphi survey) and assertion date. Assertion criteria for gene-disease validity classifications are also required. Submitters can provide optional information such as comments about the curation, PubMed identifiers, or a link to a public display of the curation in their database.

### Submission Validation

Submission sheets provided by GenCC submitters go through an automated validation process which checks to confirm the submitted IDs (HGNC ID, OMIM Phenotype MIM number, ORPHAcode, MONDO ID, HPO ID, GenCC Classification ID, and GenCC submitter ID) are valid identifiers. This is done using data provided by HGNC and the Monarch Disease Ontology API. Submission sheets with validation discrepancies are reviewed manually by staff in conjunction with the submitter to resolve submission processing errors. Submission sheets that pass validation are processed and displayed in a staging environment where staff are able to review and spot-check submission records. Once approved the submissions are published to the publicly available search interface.

### Browser and Downloads

Gene-disease validity assertions are available through a searchable browser interface as well as through downloadable formats (XLSX, XLS, TSV, CVS). The data available in the download include a GenCC unique submission ID; gene ID and symbol; disease ID and label; classification ID and label; Mode of Inheritance ID and label; Submitter ID and label; public report URL; submission notes; URL for assertion criteria; submission PMIDs; and dates related to the submission. For updating submissions, the submission file also includes fields for the original IDs and labels included in the original submission.

## Results

### Comparison of gene-disease curations across efforts

When GenCC member groups first met, each presented an overview of their resource, methods used for determining gene-disease validity, and terms used to describe the strength of a gene-disease relationship (Table S2). Some groups used terms to describe the likelihood of a relationship, while others used terms to describe the strength of evidence, and others used colors to indicate whether a gene should be added to a disease-specific gene panel. Viewing these terms side by side, members agreed that in order to create a database of gene-disease assertions, harmonization of terms would need to be completed and thus, a modified Delphi survey process was performed.

### Delphi Survey

A Delphi survey was developed to decide on harmonized terms for the description of the validity of a gene-disease relationship in the context of a monogenic disease. Future efforts will be made to tackle complex disease and those conditions that display low penetrance. In general, survey takers across all three rounds were relatively familiar with genetics; on a scale of one (unfamiliar) to five (very familiar), 205 individuals rated at a five, 31 at a four, and five at a three, with zero respondents rating a two or a one. When asked to describe their profession with multiple responses allowed, the top three answers were “Researcher” (100 responses) followed closely by “Clinical Genetics Laboratory Director or Staff” (96 responses) followed by “Medical Genetics Physician” (62 responses).

The survey was completed in three rounds (Methods and Figure 1). In total, 12 evidence and 27 likelihood terms across seven gene-disease validity groups were proposed while drafting the survey which was then taken in round 1 by GenCC members regularly participating in monthly calls (N=33). In round 2, extended GenCC membership, such as curators from GenCC groups, took the survey (N=38). After round 2, calculations detailed in the methods were used to narrow down the possible terms. Sixteen terms fell two or more standard deviations below the mean for each gene-disease validity definition and were eliminated (Figure S1), for example *verified, feasible, promising*, and *unconvincing*. The final round of the survey was sent to the following groups which had responses as noted: ESHG (59 responses), ASHG/CCMG (51), ClinGen (38), ACGS/BSGM (38), AMP (28), AGNC (9), Australian Genomics (6), users of PanelApp (4), and was posted on Twitter (8) for a total of 241 total responses received. The number of views of the optional introductory video was 320 as of 1/15/19 when the survey was closed, which suggests that a high percentage of individuals watched it before taking the survey. The final validity terms chosen by the genetics community (Table 1) were 93% concordant with the GenCC membership choices, and all member groups agreed to adopt the new consensus terms for use within the GenCC database and consortium activities. Some groups planned to move their curation systems to conform to the new categories and terms as soon as possible and others chose to map their existing terms to the consensus terms. The “evidence terms” set was chosen as the terms to use for database display and has become the primary term set used across the GenCC. The exception is for Orphanet and OMIM, two well-established public resources, which are mapping to the less granular term *supportive*, given that these resources do not distinguish between Limited, Moderate, Strong and Definitive.

**Table 1.**
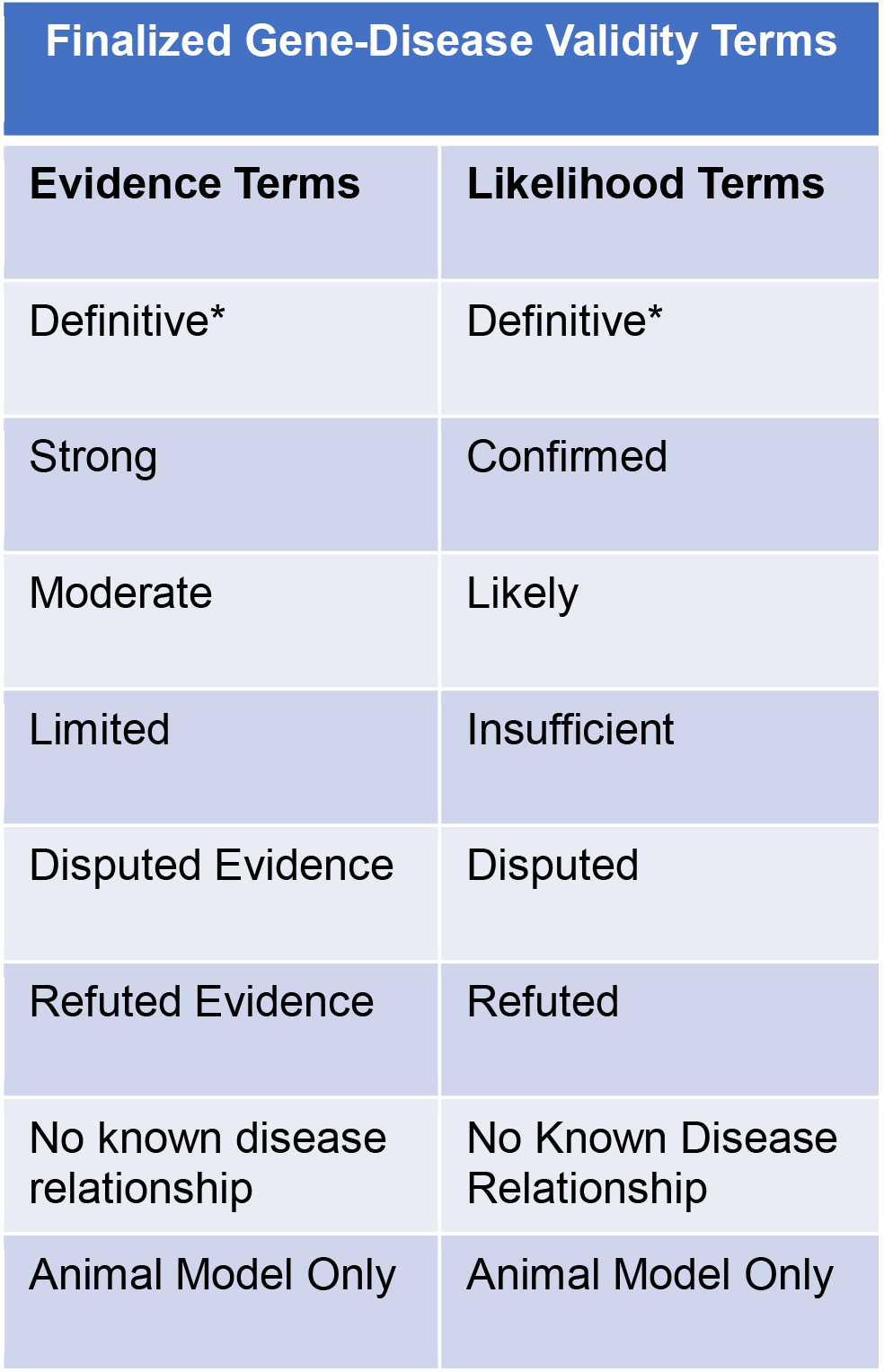
Finalized gene-disease validity terms from the modified Delphi survey. The “evidence terms” set was chosen for the database display and has become the primary term set used across the GenCC. *Definitive was not surveyed as a separate term choice. Definitions for each term can be found in the supplement (Table S1)

### GenCC Database

The GenCC database beta version was launched in December 2020. Between January 1 and September 10, 2021 there were 36,470 page views by 4,126 users. Website views of the database by country are presented in Figure 2a. The top seven countries with database users are: United States (1300), China (826), United Kingdom (375), Germany (148), Australia (139), Israel (127), and Canada (120) (Figure 2b).

**Figure 2:**
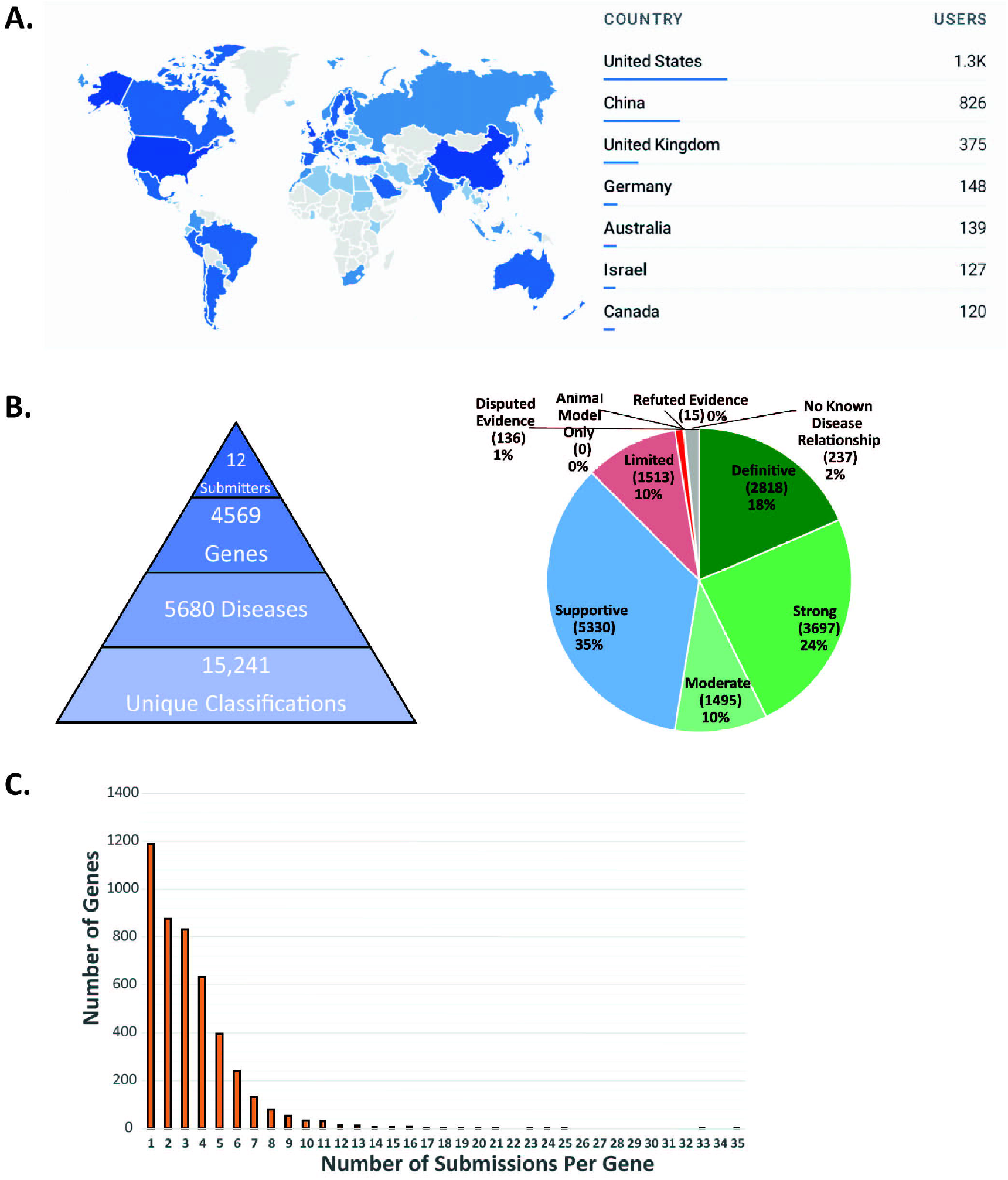
Summary statistics for the GenCC database. A) Map of users by country with the top 7 countries listed as of 9/14/21. B) A summary of all data submitted to the GenCC database as of 12/21 including a breakdown of gene-disease validity claims. C) A graph of the number of submissions per gene (N=4569 genes, average is 2 submissions)

The database currently contains 15,241 submitted classifications on 4,569 unique genes from 12 submitters (Figure 2b). All submitted assertions are harmonized to the evidence terms decided upon in the modified Delphi survey. As of December 2021, there were 2,818 Definitive, 3,697 Strong, 1,495 Moderate, 1,513 Limited, 136 Disputed Evidence, 15 Refuted Evidence, 237 No Known Disease Relationship, 0 Animal model only, and 5,330 Supportive assertions (Figure 2b). Similar to Orphanet submissions, “Supportive” will be applied to OMIM data pending availability of an API-based submission process. The average number of unique submissions per gene is two with the maximum for any entry being 35 (Figure 2c). The average number of unique submitters per gene is two with the maximum number for any entry being seven.

There are multiple different views of the database. Each will be described briefly here, but for more information please refer to our FAQ page on the website (https://thegencc.org/faq#website-pages-faq). The database landing page (Figure 3a) lists all curated genes. Each line corresponds to one unique gene which is collapsed, but can be expanded to view a summary of all of the submissions for that gene. Genes are displayed and searchable using the current HGNC gene symbol. Additionally, HGNC, a GenCC member group, is committed to stabilizing the gene symbols of all of those genes curated by the GenCC. Those genes with stabilized symbols have a “stable” tag on the HGNC website. Please refer to the HGNC website for more information (https://www.genenames.org/). GenCC users can filter by HGNC gene symbol, disease, or submitter at the top of the landing page. The harmonized evidence terms each correspond to a particular color and are listed at the top of the screen as filterable check boxes. Clicking on the details button for a gene will take users to the gene-specific classification page. There are multiple tabs on the gene-specific page that display the entries by classification (Fig 3b), disease, and submitter. All of these pages display: the harmonized gene-disease validity term, the HGNC gene symbol, the submitted disease (accepted ontologies are OMIM, Orphanet, and MONDO), the mode of inheritance (using HPO terms), the evaluated and submitted dates, the submitter, links to assertion criteria, and any public curation report on a submitter’s own website. For grouping purposes, all diseases are mapped to MONDO, which contains entries for all OMIM and Orphanet diseases. These grouped diseases are called “disease equivalents’’. If a user clicks “more details” for a particular entry on a disease page, this displays a submitter-specific page that often includes PMIDs or additional evidence related to the assertion (Figure 3c). The submitter pages include a brief description of the resource, a website link, a contact person, summary statistics for that submitter, and a public link to their assertion criteria for gene-disease validity classification (Figure 3d). The statistics page includes summary statistics (similar to Figure 2). The downloads page provides multiple formats for database downloads (see methods for more information).

**Figure 3:**
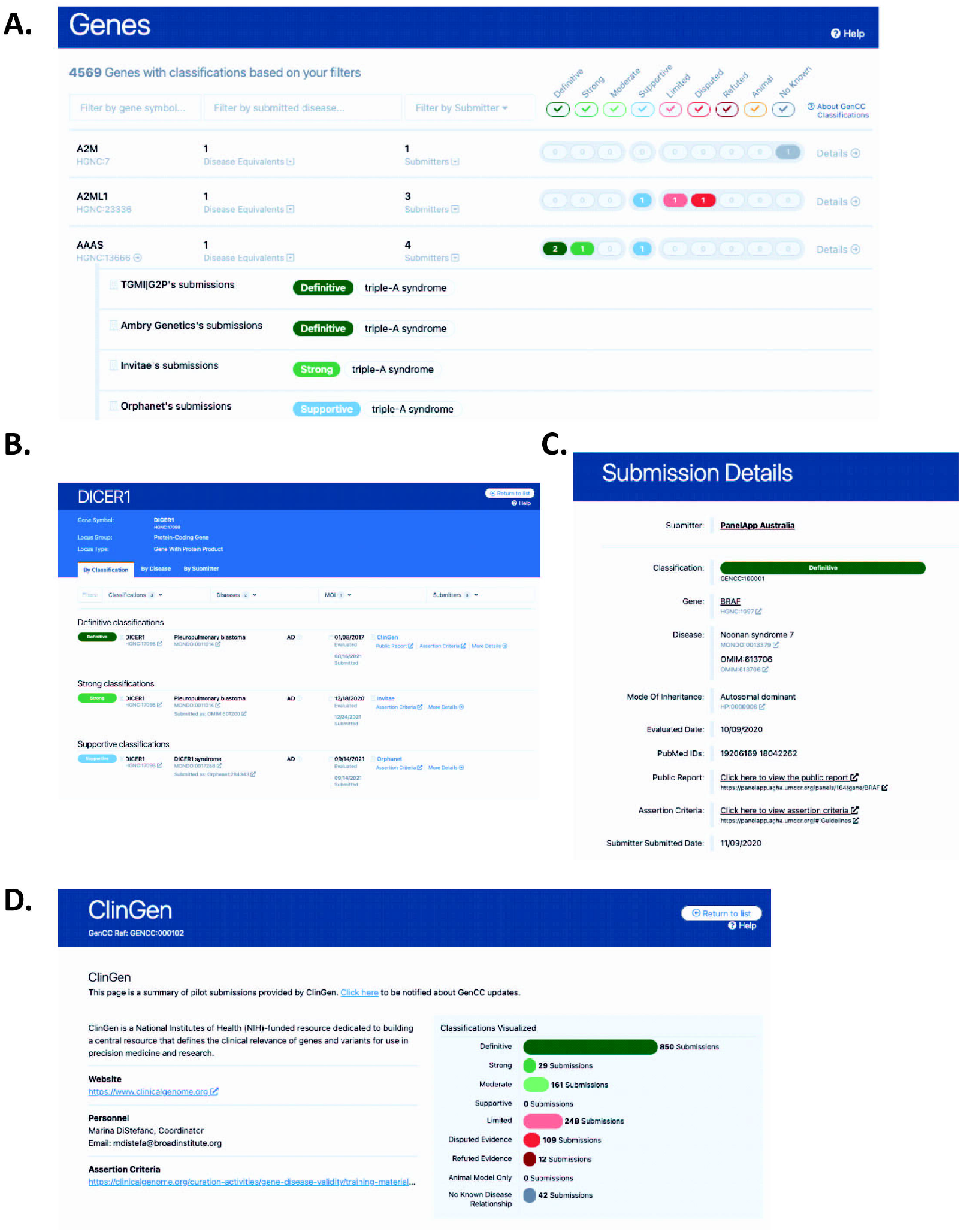
Screenshots of the GenCC database: A) the landing page; B) the gene level page sorted by classification C) Submission-specific page for an entry; D) Submitter page

### Discrepancy Resolution

Studies have demonstrated that interpretation differences can be resolved with data sharing.^6-9,11,13,14^ With the launch of the GenCC database, we plan to support the resolution of differences in gene-disease assertions in an evidence-based manner, by analyzing conflicts in the database and sharing evidence amongst GenCC submitters. Despite the use of an ontology, this effort often requires manual review because of the general lack of harmonization of disease terms. Although submissions are mapped to MONDO, submitters will often use different levels in a disease ontology or disease name synonyms that are not calculated to be exact matches. For instance, one submitter may assert a limited role in ‘breast cancer’ for *BRIP1* and another may assert a definitive role in ‘breast and ovarian cancer’ for *BRIP1* and these claims may both be valid given that *BRIP1* is validly implicated in ovarian cancer but not breast cancer.^17-19^ To begin these efforts, we have calculated some gene-disease validity term conflict statistics from the database. For this analysis (Table 2), a conflict was defined as a Limited/Disputed/Refuted assertion versus a Moderate/Strong/Definitive assertion. Three types of conflicts were calculated: 1) “Level 1 conflicts” were those conflicting entries where the gene matched, without regard to the mode of inheritance or disease term, but gene-disease validity terms conflicted: 13.4% (610) of all submitted genes had at least one such conflict; 2) “Level 2 conflicts” were those conflicting entries where gene and mode of inheritance matched but gene-disease validity terms conflicted. These were agnostic of submitted disease terms. 10.7% (488) of all genes had at least one such conflict; 3) “Level 3 conflicts” were those where gene, disease term, and mode of inheritance all matched but gene-disease validity terms conflicted: 5.4% (246) of all submitted genes had at least one such conflict.

**Table 2.** Conflict analysis of GenCC submissions. For the purposes of conflict analysis, a conflict is defined as a Limited/Disputed/Refuted assertion vs a Moderate/Strong/Definitive assertion. Counts and percentages represent conflicting genes. Conflict analysis was performed for 4569 genes on 12/2021. AD, Autosomal Dominant; AR, Autosomal Recessive; MOI, Mode of Inheritance

| Conflict Type | Number of Genes (%) | Examples |
| --- | --- | --- |
| **Level 1 conflicts:** Gene matches (agnostic of MOI and Disease term) | 610 (13.4%) | Submitter 1:TNNT3, distal arthrogryposis type 2B2, AD, Moderate<br>Submitter 2:TNNT3, nemaline myopathy, AR Limited |
| **Level 2 conflicts:** Gene and MOI matches (agnostic of Disease term) | 488 (10.7%) | Submitter 1: ADAMTS18, microcornea-myopic chorioretinal atrophy-telecanthus syndrome, AR, Strong<br>Submitter 2: ADAMTS18, inherited retinal dystrophy, AR, Limited |
| **Level 3 Conflicts:** (Gene, Disease term, MOI all match) | 246 (5.4%) | Submitter 1: ABAT, GABA aminotransferase deficiency, AR, Strong<br>Submitter 2: ABAT, GABA aminotransferase deficiency, AR, Limited |

## Discussion

With the increased use of exome or genome analysis, as well as reliance on disease-focused gene panels for genetic testing and genomic analysis, confidence in the validity of a gene-disease association is more important than ever before. The GenCC was formed to help standardize terms surrounding gene-disease validity curation and share gene curation data publicly. To that end, we performed a modified Delphi survey to standardize terms describing gene-disease validity. GenCC members created standardized definitions and terms were chosen after three rounds of surveys. In the final round, the genetics community was surveyed. The finalized terms that were chosen are now used for submission to the GenCC database.

Launched in December 2020, the GenCC database is, conceptually, a similar resource to ClinVar, but for gene-disease assertions instead of variant-disease assertions. Currently, the database has 15,241 submitted classifications on 4,569 unique genes from 12 submitters. GenCC members submit assertions to this database and total database content is downloadable for use in clinical annotation pipelines and research activities. Although database growth is driven by submissions, we anticipate that it will be updated bimonthly until API-based submissions and data access is supported. Both DECIPHER^15^ and the UCSC genome browser^16^ currently display GenCC curations in their resources and both ClinGen and HGNC provide links to GenCC for each gene displayed on their websites. Planned future enhancements for the database include an API for both submission and database use. OMIM plans to begin contributing once the API is available to connect its large dataset to the GenCC database in real time. Additional enhancements include user-specific search parameters and a panel builder function that allows searching by keyword, Human Phenotype Ontology terms,^20^ or any disease term or identifier present in a MONDO hierarchy to generate a filterable gene list. The list could then be downloaded and used to inform clinical testing panel design or indication-based analysis during exome or genome sequencing.These enhancements will help to harmonize gene-disease validity across clinical and research curation efforts.

GenCC will begin to resolve gene-disease validity discrepancies across GenCC submitters using a manual review process where simpler discrepancies (e.g. one submitter’s curation is out of date) will be handled offline by respective submitters and complicated discrepancies will be discussed on our monthly conference calls. We will also engage ClinGen Gene Curation Expert Panels for discrepancy resolution where they exist for the relevant disease area. Terminology standardization, sharing of gene-disease validity classifications and resolution of curation conflicts will facilitate collaboration across international curation efforts and in turn, improve consistency in genetic testing and variant interpretation. We welcome participation from additional groups performing evidence-based curation of gene-disease validity.

## Supporting information

All supplemental tables and files

GenCC data snapshot 12_2021

## Data Availability

All data from the GenCC website is openly available in multiple
download formats and can be accessed here (https://search.thegencc.org/download). A
snapshot of the GenCC database (Dec 2021) relevant to the figures and analysis in this
manuscript can be found in the supplemental files. Deidentified Delphi survey
responses are available upon request.

## Data Availability

All data from the GenCC website is openly available in multiple download formats and can be accessed here (https://search.thegencc.org/download). A snapshot of the GenCC database (Dec 2021) relevant to the figures and analysis in this manuscript can be found in the supplemental files. Deidentified Delphi survey responses are available upon request.

## Acknowledgements

Support for title page creation and format was provided by AuthorArranger, a tool developed at the National Cancer Institute. This study was supported by the National Human Genome Research Institute of the National Institutes of Health under award U24HG006834. The content is solely the responsibility of the authors and does not necessarily represent the official views of the National Institutes of Health or other affiliations. This work was supported by the Intramural Research Program at the National Library of Medicine. PanelApp Australia is supported by Australian Genomics (NHMRC Grants GNT1113531 and GNT2000001). This work was supported by Wellcome Trust [107469/Z/15/Z; 200990/A/16/Z], Medical Research Council (UK), British Heart Foundation [RE/18/4/34215], the NIHR Imperial College Biomedical Research Centre. We thank all PanelApp reviewers and those who have contributed feedback or gene lists to help in the development of PanelApp; individual panels show the names and affiliations of contributors. We thank all participants in the 100,000 Genomes Project. This research was made possible through access to the data and findings generated by the 100,000 Genomes Project. The 100,000 Genomes Project is managed by Genomics England Limited (a wholly owned company of the Department of Health). The 100,000 Genomes Project is funded by the NIHR and NHSE. The Wellcome Trust, Cancer Research UK and the Medical Research Council have also funded research infrastructure. The 100,000 Genomes Project uses data provided by patients and collected by the NHSE as part of their care and support. Open Targets is supported by Open Targets. The work performed by authors at EMBL-EBI was supported by the Wellcome Trust [WT200990/Z/16/Z]. For the purpose of open access, the authors have applied a CC BY public copyright licence to any Author Accepted Manuscript version arising from this submission.

## Author Contributions

Conceptualization: M.T.D.; J.A.; J.S.B.; E.B.; E.A.B.; H.C.; F.C.; L.C.D.; H.V.F.; D.R.F.; R.E.F.; J.G.; A.H.; M.R.H.; S.E.L.; C.L.M.; E.M.M.; A.O.; E.M.R; E.R.; A.M.R.; Z.S.; S.T.; J.S.W.; C.F.W.; H.L.R; Data curation: M.T.D.; S.G.; F.S.A.; J.A.; M.A.; C.A.; M.B.; C.B.; A.J.C; L.C.D.; Y.E.; R.E.F.; S.E.L.; I.US.L.; S.M.; E.M.M.; A.O.; A.P.; K.R.; A.R.; C.R.; C.S.; Z.S.; J.T.; E.W.; T.M.Y.; Formal analysis: M.T.D.; S.G.; L.B.; P.W.; H.L.R; Methodology: M.T.D.; S.G.; H.L.R.; Project administration: M.T.D.; Software: S.G.; P.W.; Supervision: M.T.D.;C.L.M.; H.L.R; Validation: M.T.D.; S.G.; L.B.; P.W.; Visualization: M.T.D.; S.G.; H.L.R.; Writing-original draft: M.T.D.; S.G.; P.W.; H.L.R; Writing-review & editing: M.T.D.; S.G.; F.S.A; J.A.; M.A.; C.A.; M.B.; J.S.B; E.A.B.; A.J.C.; H.C.; F.C.; L.C.D.; R.E.F.; J.G.; A.H.; S.M.; C.L.M.; E.M.M.; K.R.; E.R.; Z.S.; J.T.; S.T.; J.S.W.; P.W.; E.W.; C.F.W.; H.L.R.

