## Supplementary material for "The Gene Curation Coalition: A global effort to harmonize gene-disease evidence resources": All supplemental tables and files

Supplementary Figures:

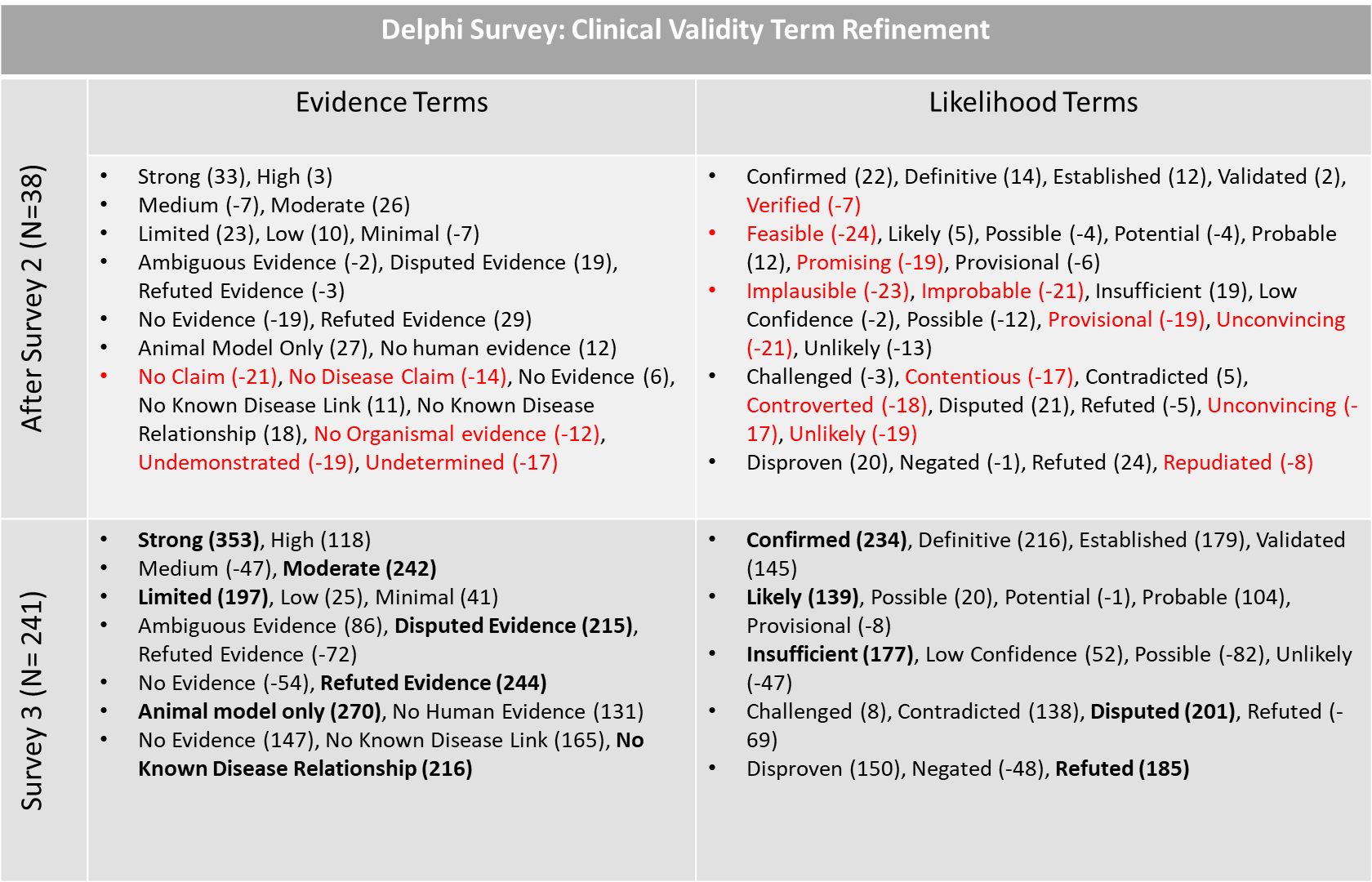

**Figure S1: Clinical Validity Term Refinement in the Delphi Survey**

Likert scale answers were converted to point values and summed (Strongly Agree= 2points, Agree = 1 point, Neutral = 0 points, Disagree = -1 points, Strongly Disagree = -2 points). Each bullet point corresponds to a different clinical validity bucket. In Survey 2, terms with scores >2 standard deviations below the average score for each category were eliminated (denoted by red text) unless there were only two term options. In survey 3, the terms with the highest scores (denoted in bold) were chosen as the final term sets.

| Finalized Term | Definition |
| --- | --- |
| Definitive* | The role of this gene in this particular disease has been repeatedly demonstrated in both the research and clinical diagnostic settings, and has been upheld over time (at least 2 independent publications over 3 years’ time). No convincing evidence has emerged that contradicts the role of the gene in the specified disease. |
| Strong (Confirmed) | The role of this gene as a monogenic cause of disease has been repeatedly and independently demonstrated, providing very convincing evidence in humans and no conflicting evidence for this gene’s role in this disease. |
| Moderate (Likely) | There is an intermediate amount of evidence in humans to support a causal role for this gene in this disease with no contradictory evidence. The body of evidence is not large (e.g. possibly only one key paper) but appears convincing enough that the gene-disease pair is likely to be validated with additional evidence in the near future. |
| Limited (Insufficient) | Little human evidence exists to support a causal role for this gene in this disease, but not all evidence has been refuted. For example, there may be a collection of rare missense variants in humans but without convincing functional impact, segregation data that could either arise by chance (e.g. across one or two meioses) or does not implicate a single gene, or functional data without direct recapitulation of the phenotype. Overall, the body of evidence does not meet contemporary criteria for claiming a valid association with disease. The majority are probably false associations. |
| Disputed Evidence | Although evidence has been reported, other evidence of equal weight challenges the claim. |
| Refuted Evidence | There has been an assertion of a gene-disease relationship in the literature, but new valid evidence has arisen that overturns the entire original body of evidence. |
| No known disease relationship | No disease claim in any organism has ever been made. |
| Animal model only | No (or very little) human disease evidence exists, but a convincing animal model exists. |

**Table S1. Definitions for gene curation categories.** Harmonized definitions for gene-disease validity levels were drafted. They are listed here alongside the finalized chosen clinical validity term for each. *Definitive was not surveyed as a separate term choice

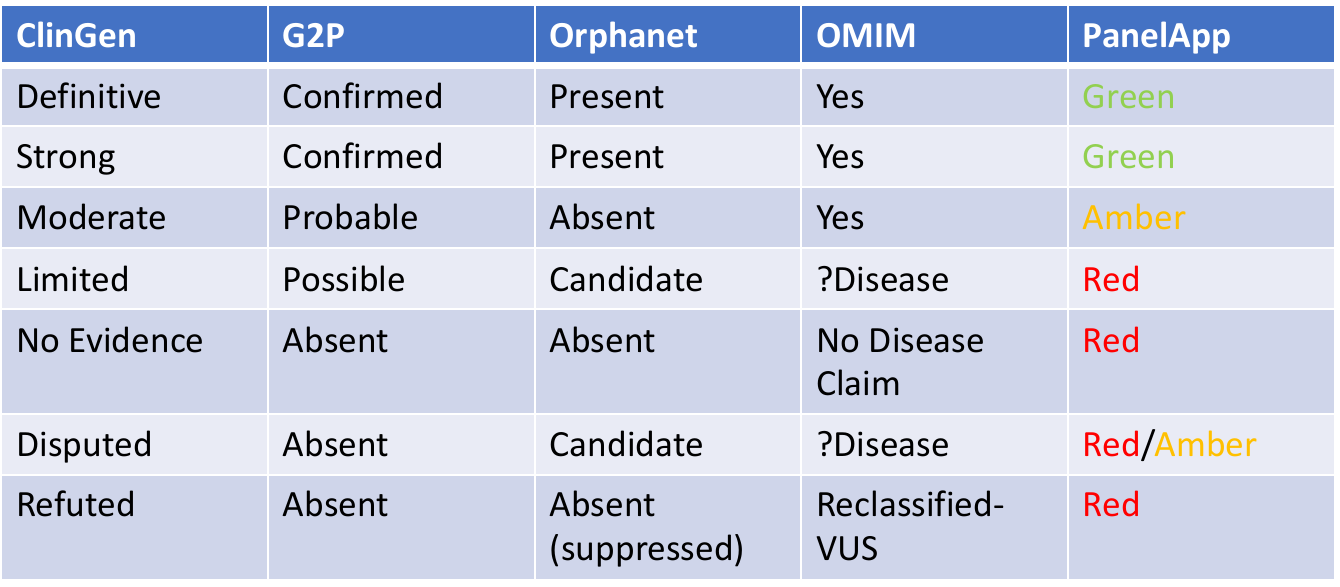

**Table S2. Clinical validity terms used by GenCC member groups before term harmonization**

**Supplementary Document: Delphi Survey Questions.** The questions of the three rounds of the modified Delphi survey are provided below.

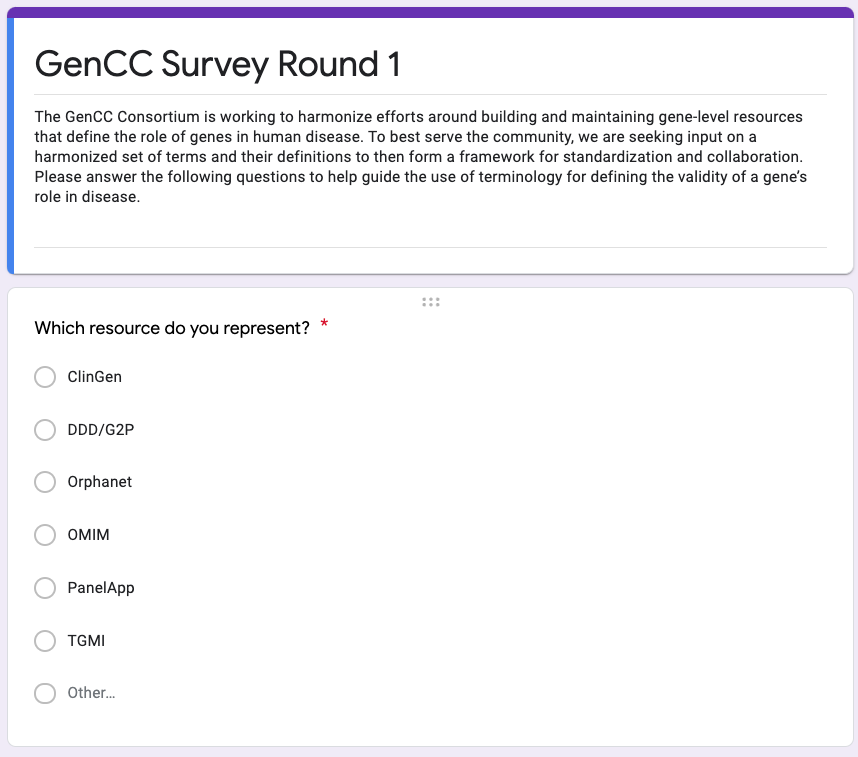

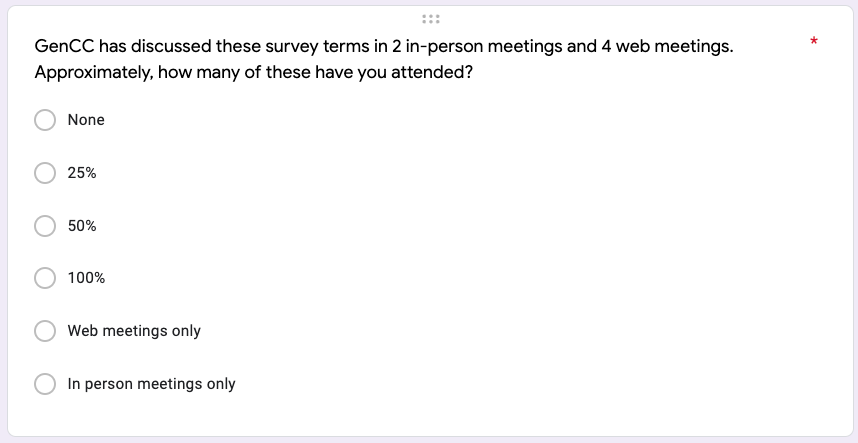

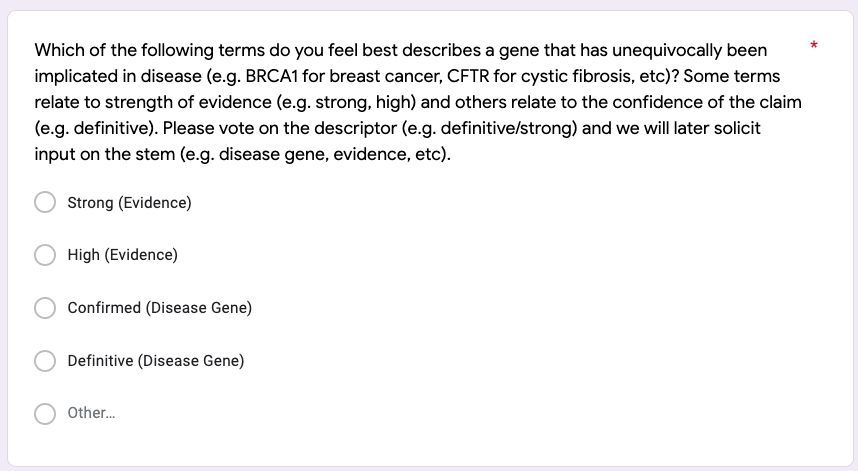

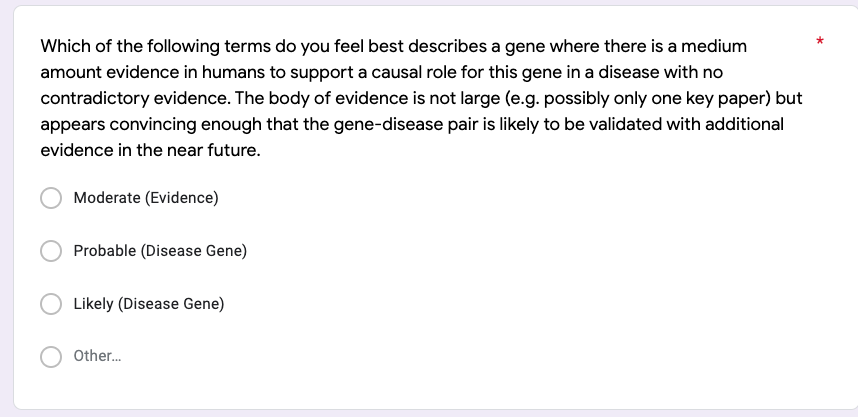

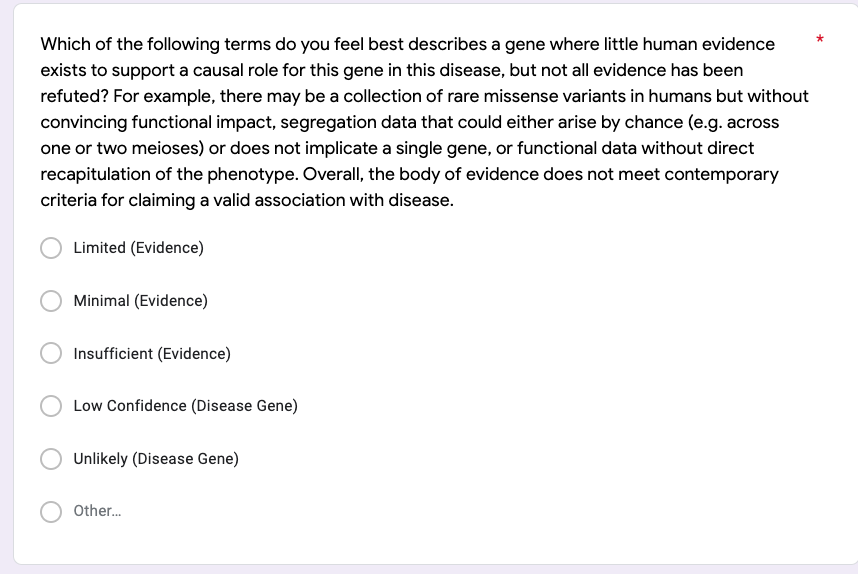

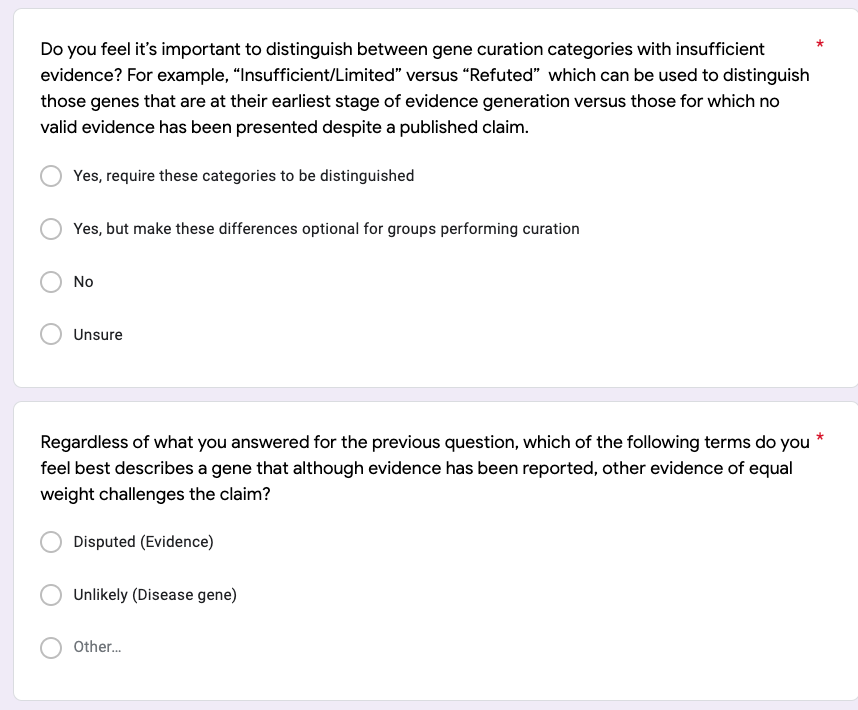

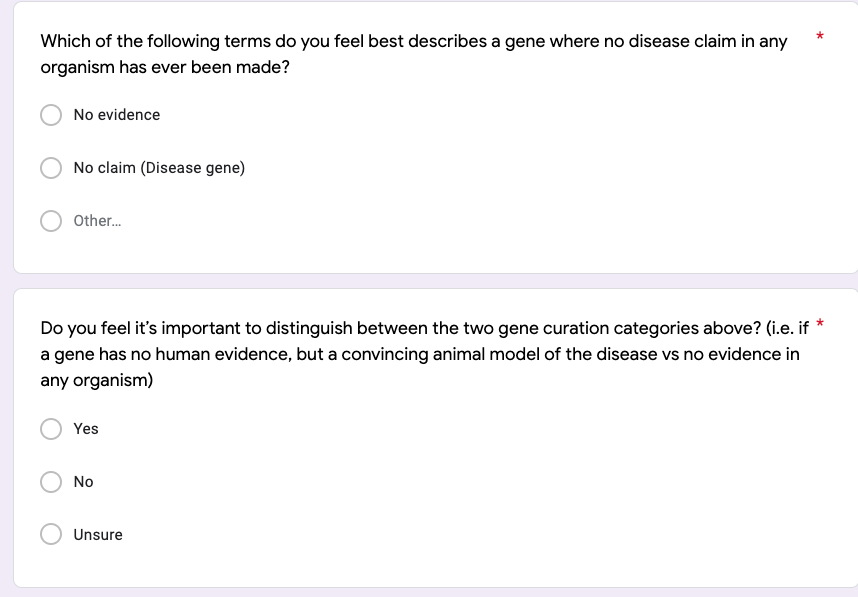

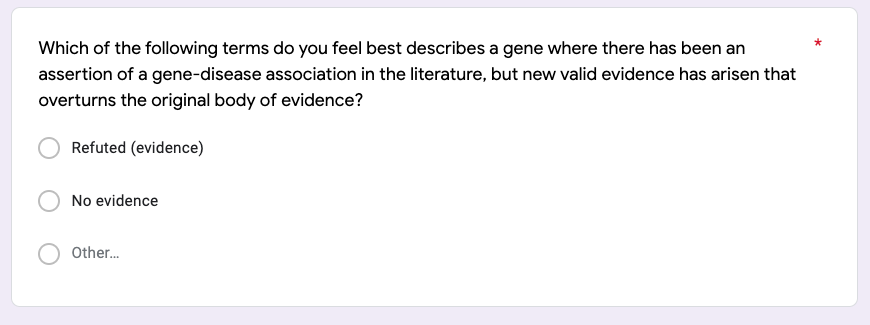

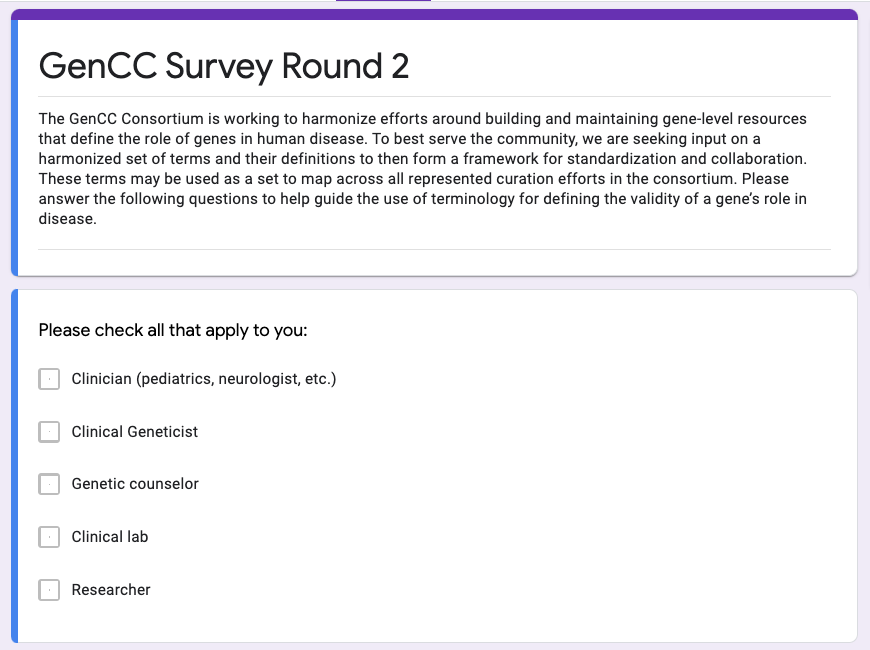

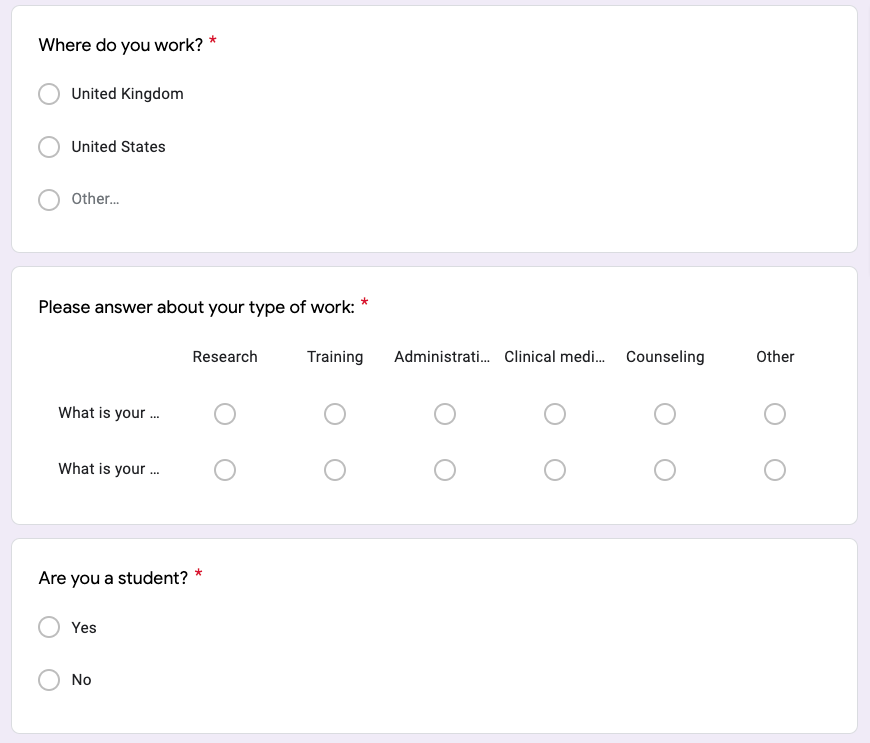

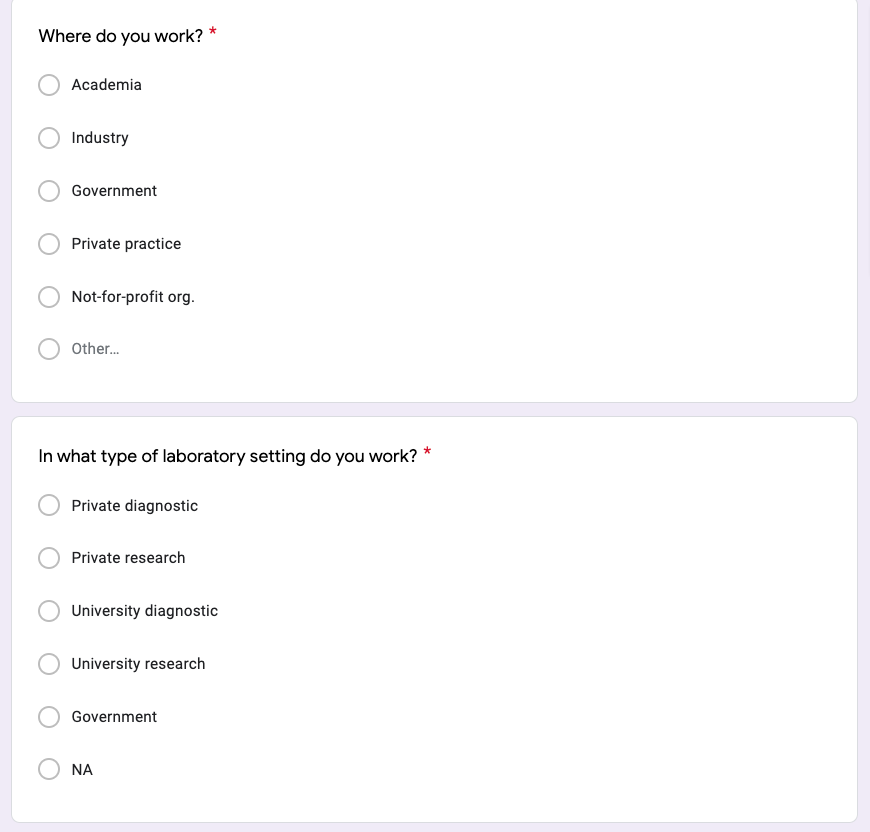

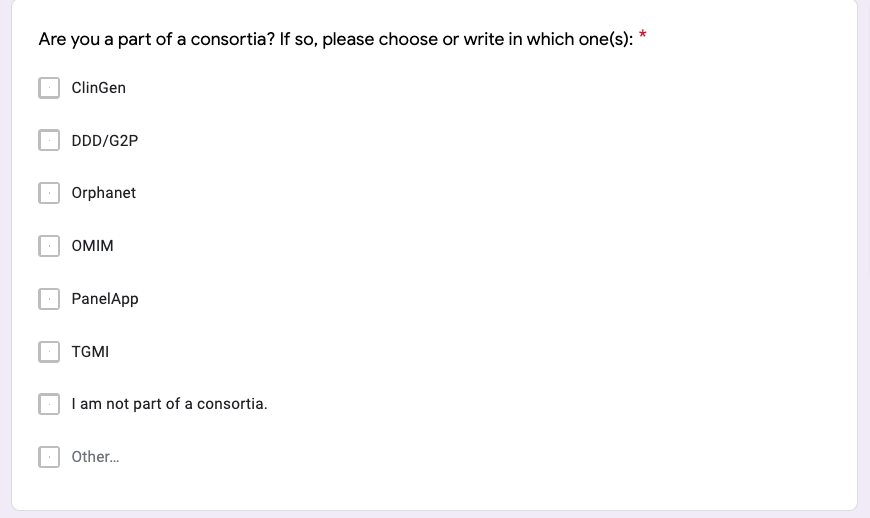

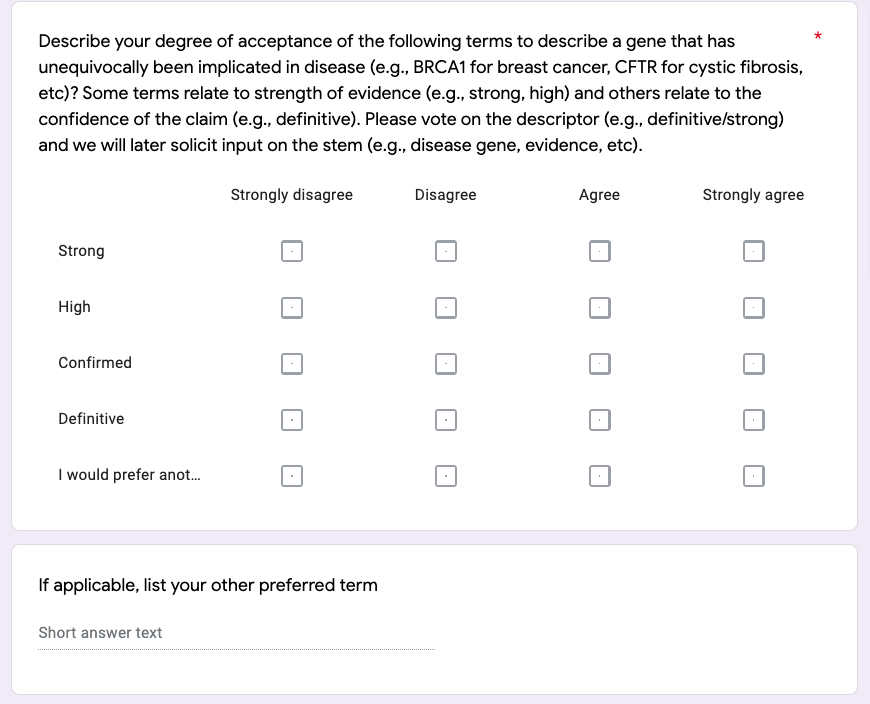

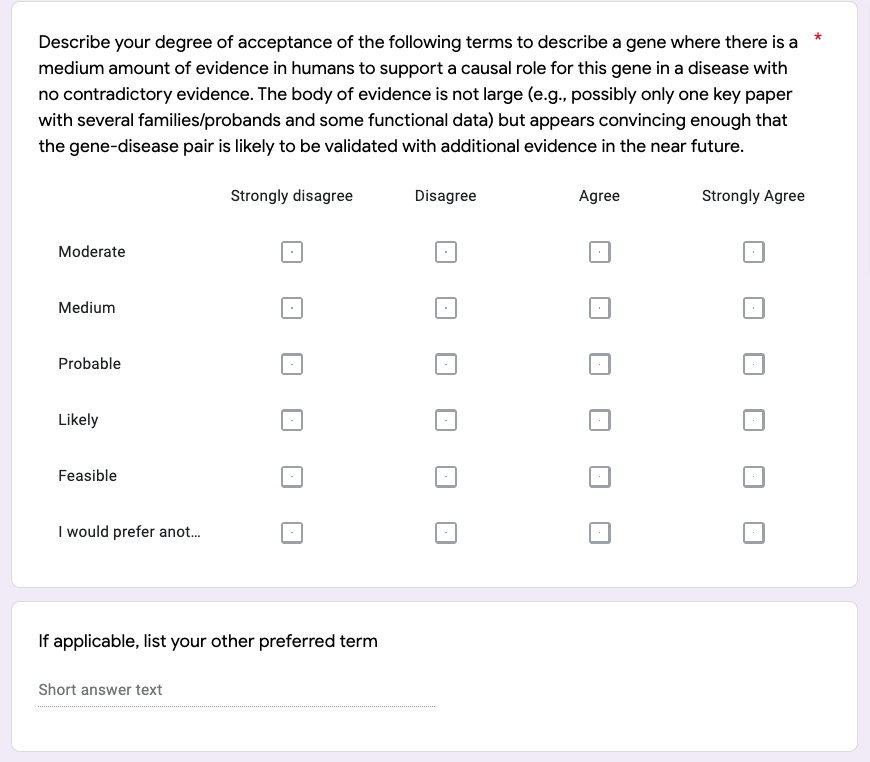

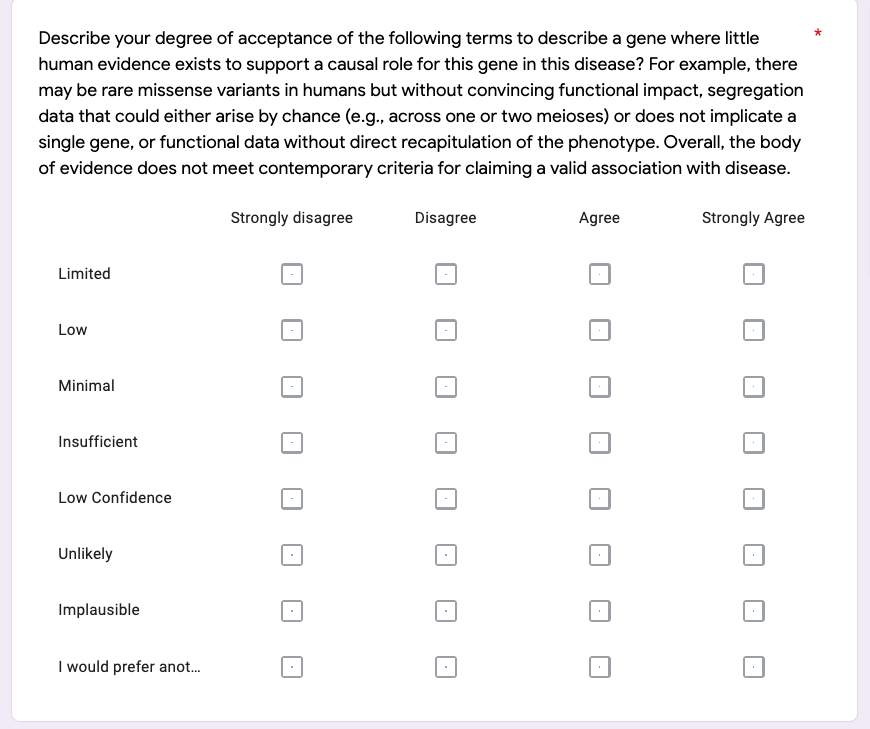

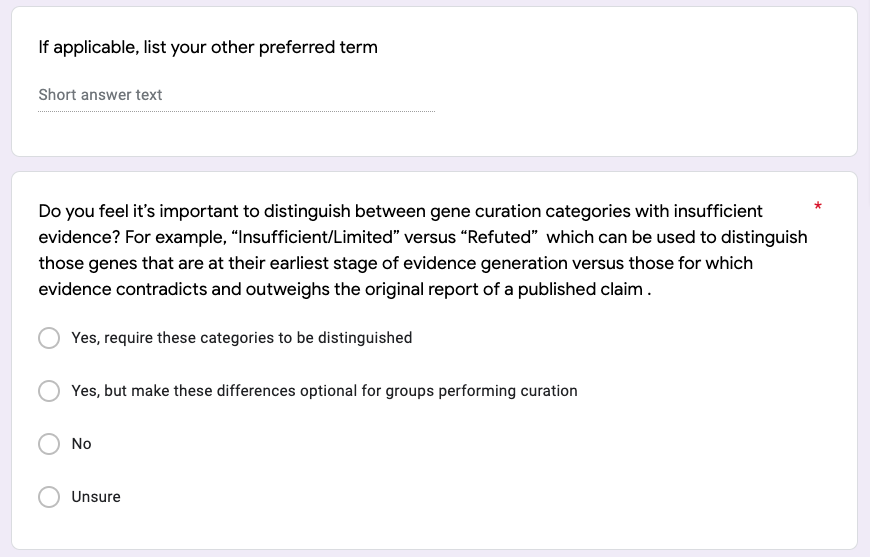

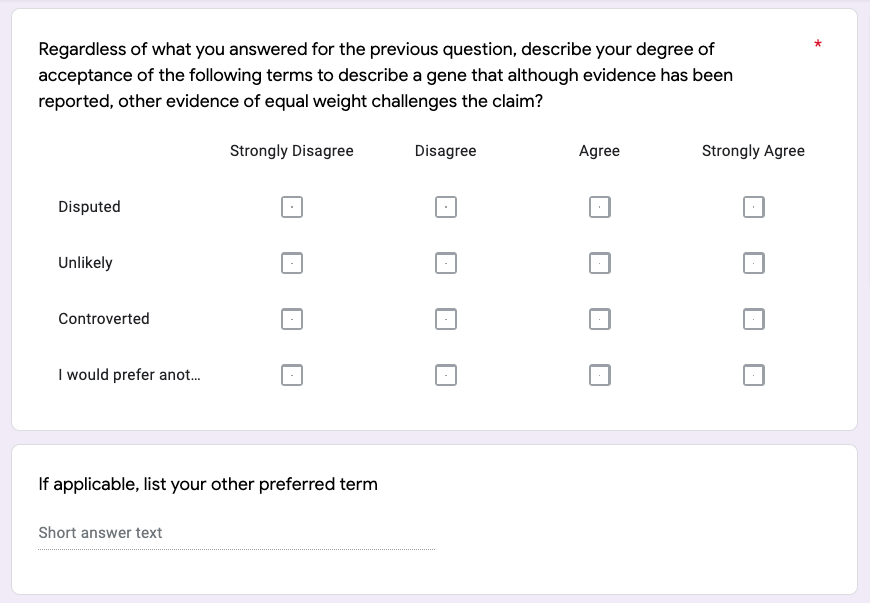

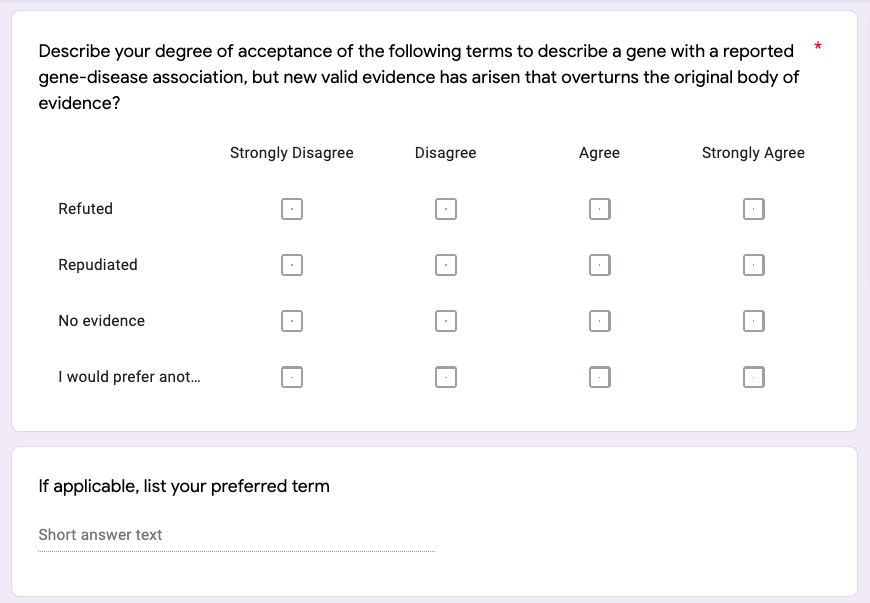

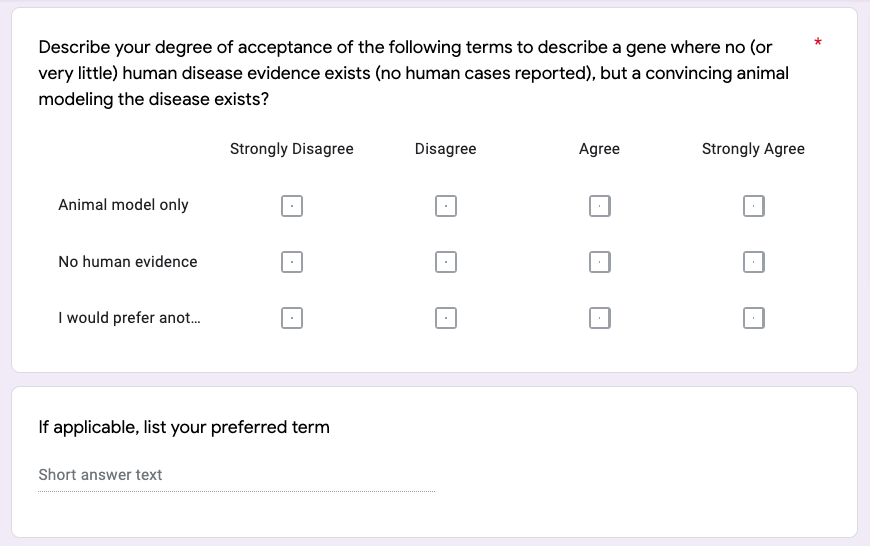

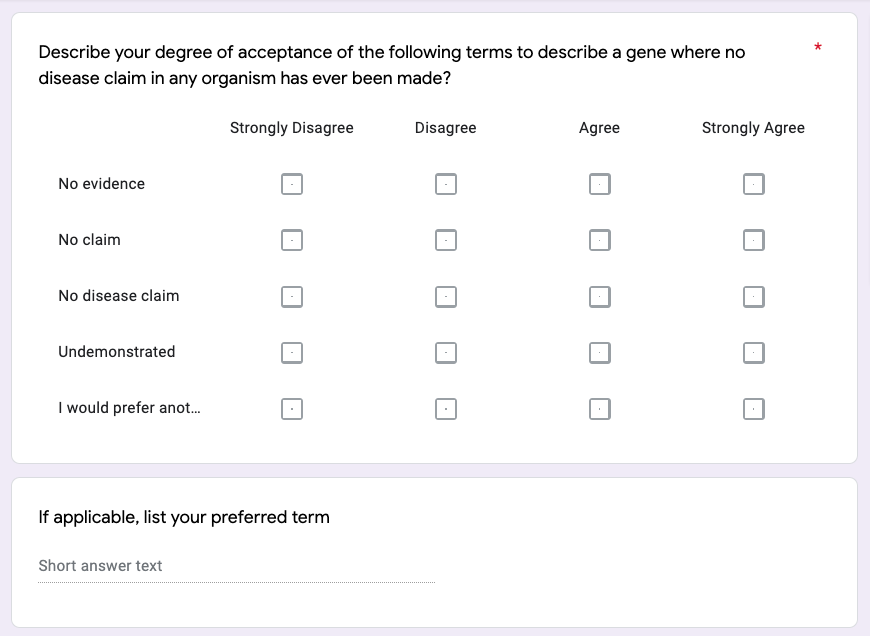

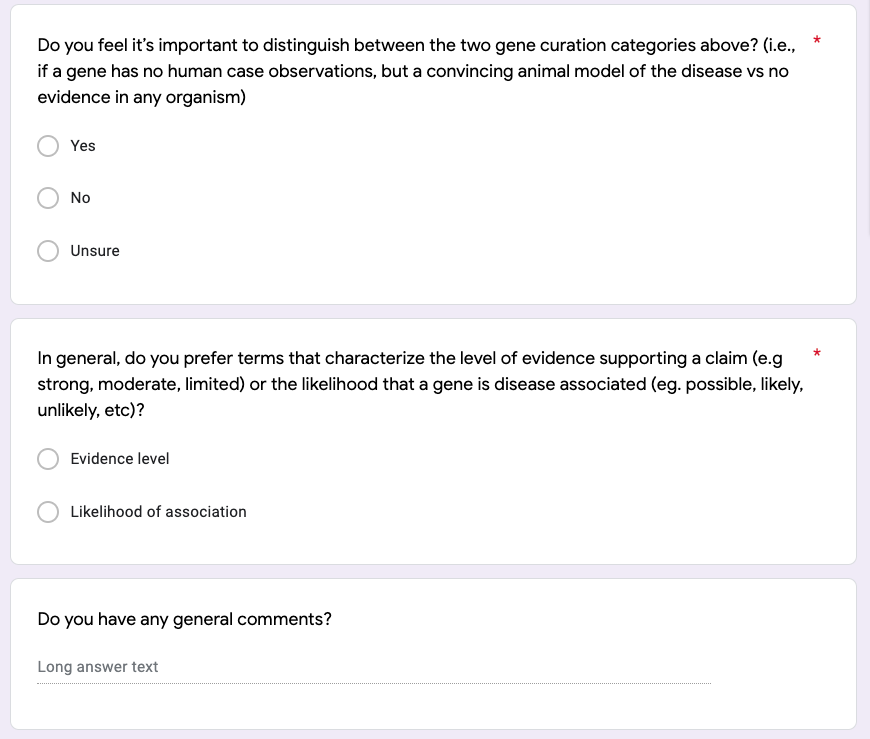

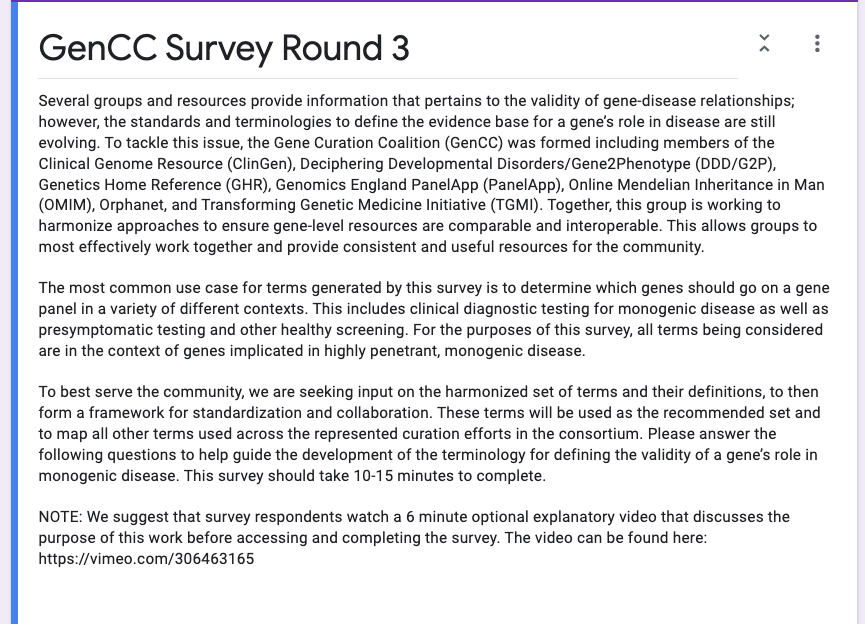

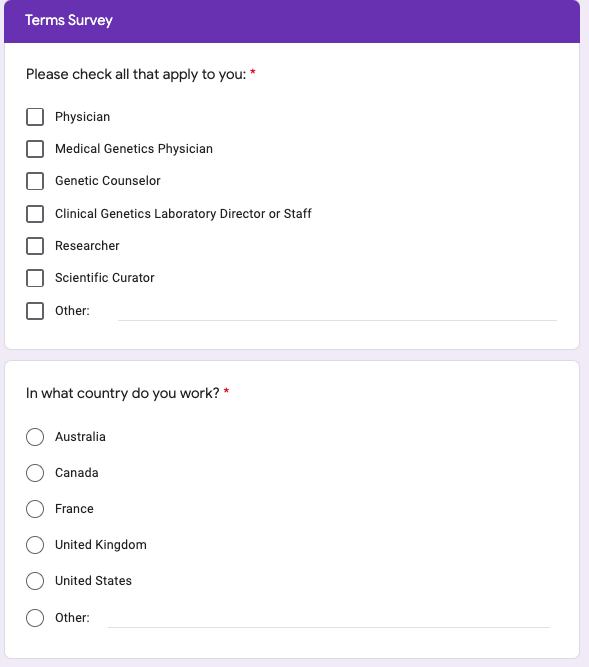

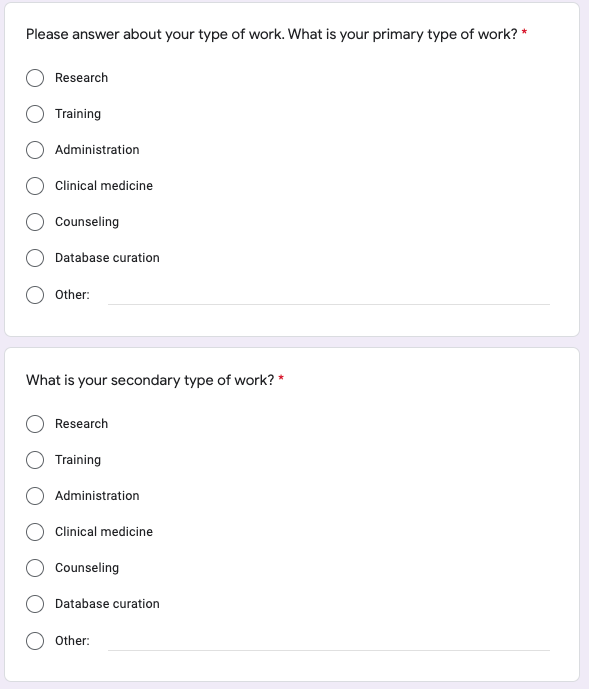

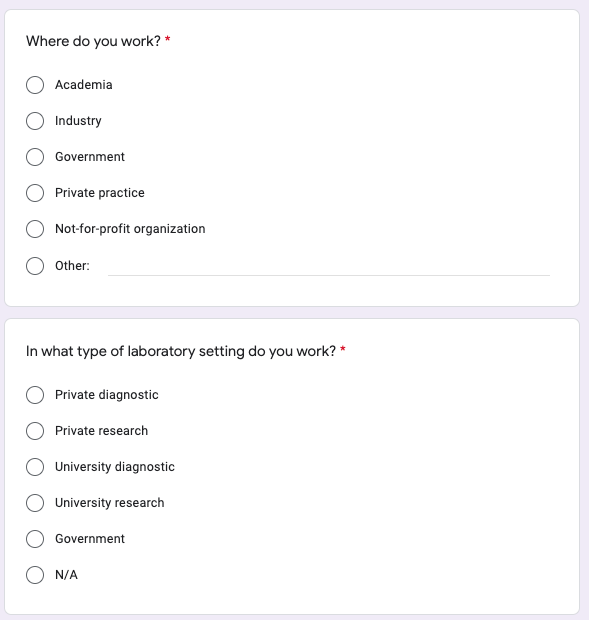

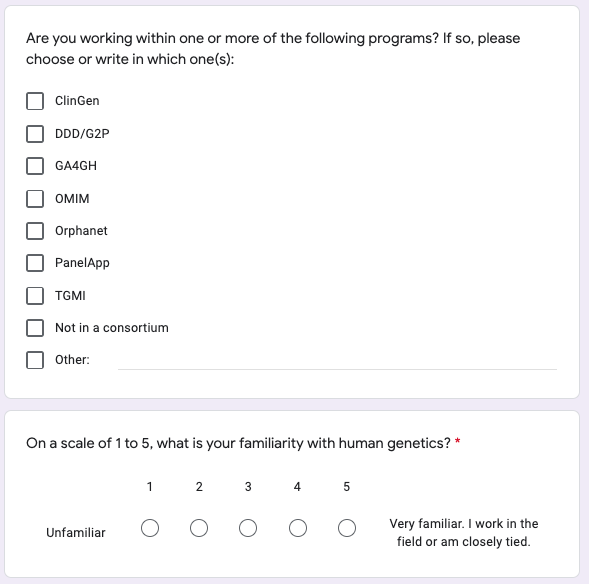

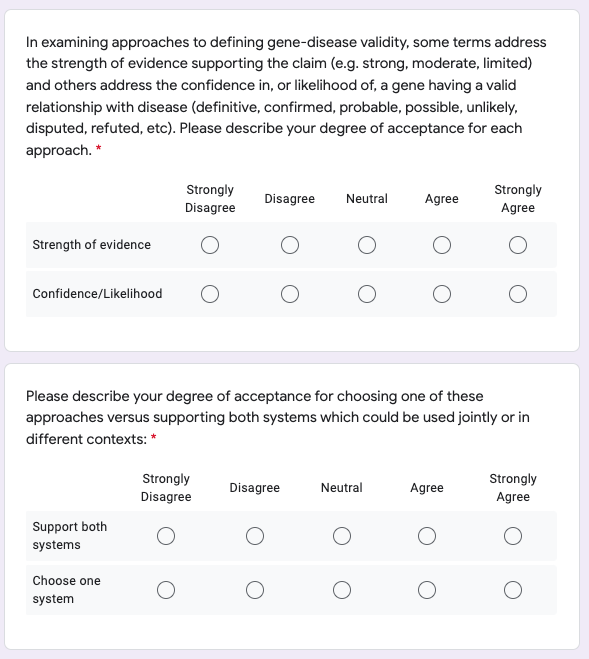

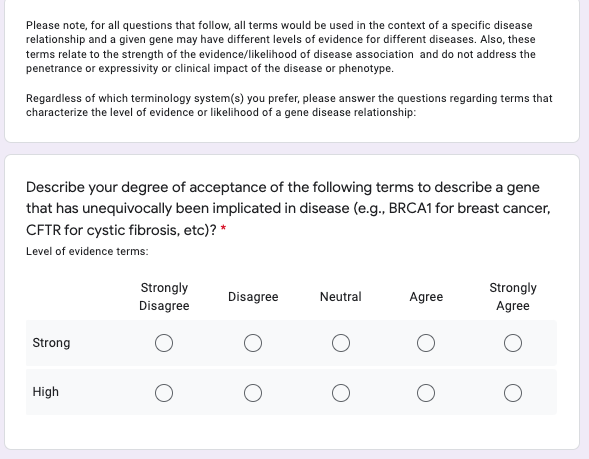

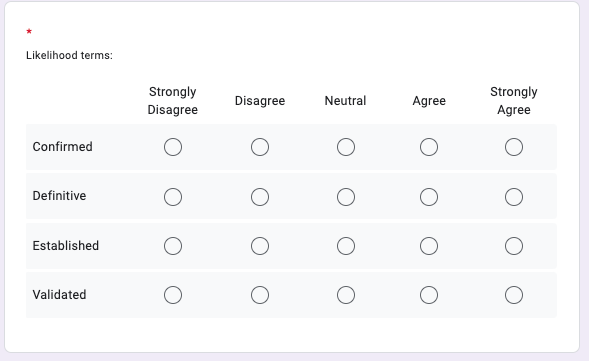

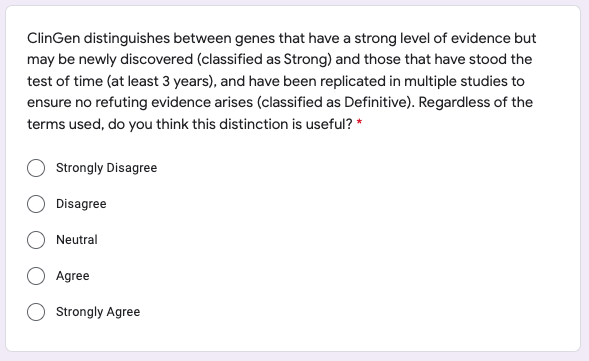

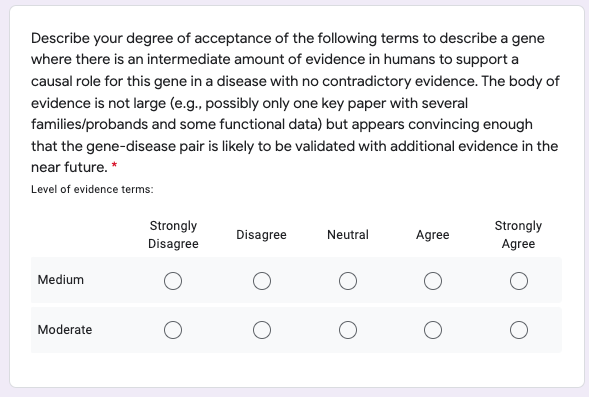

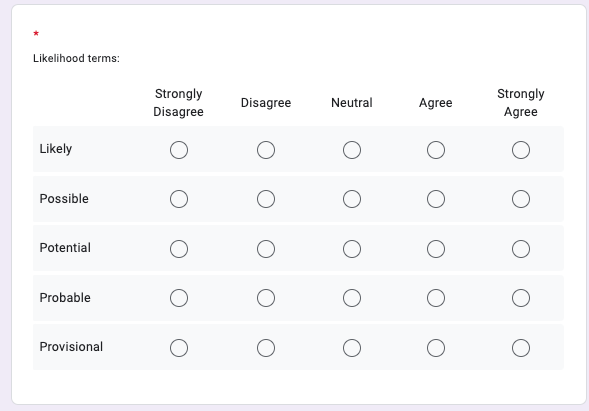

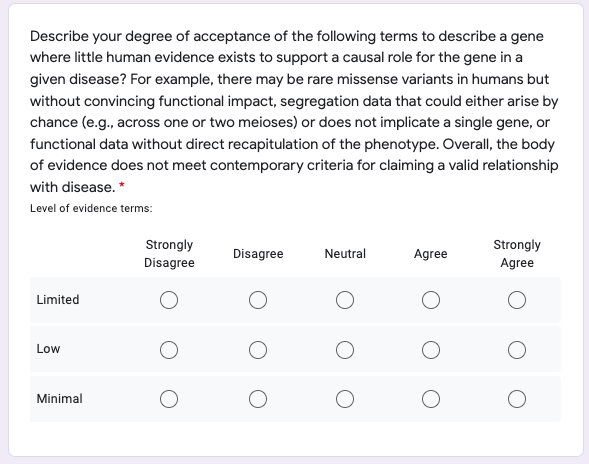

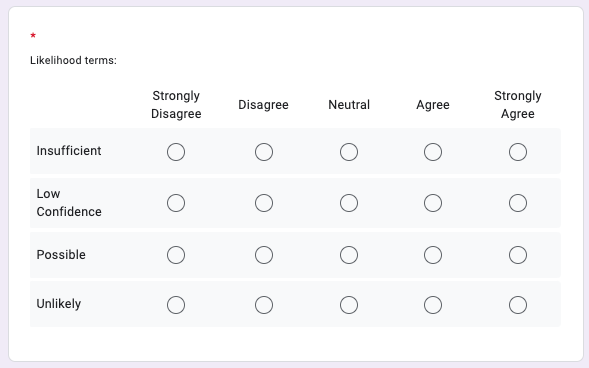

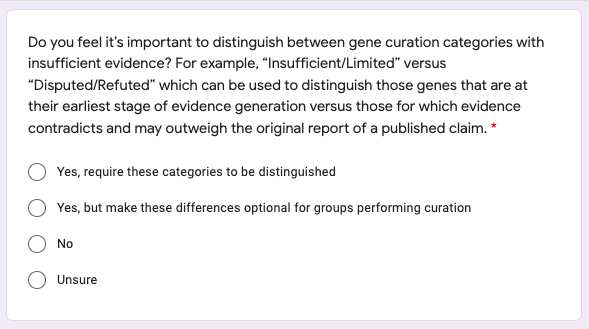

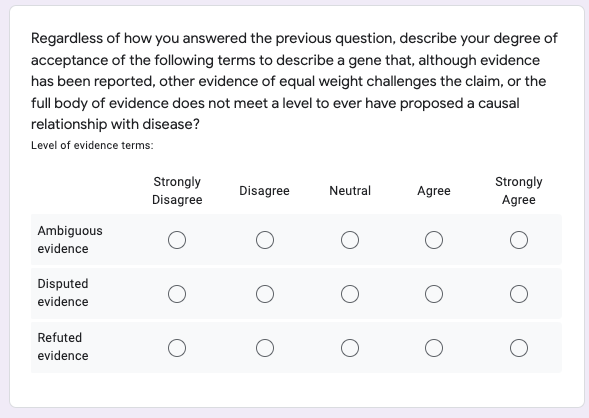

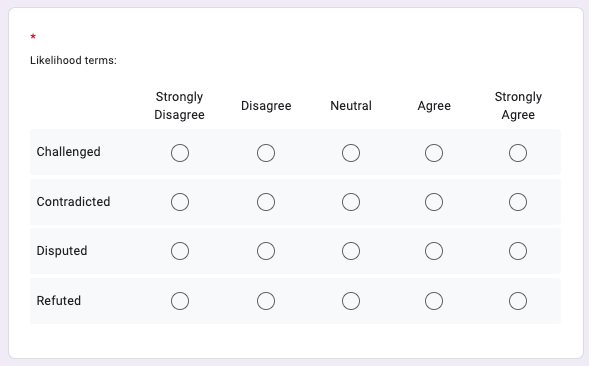

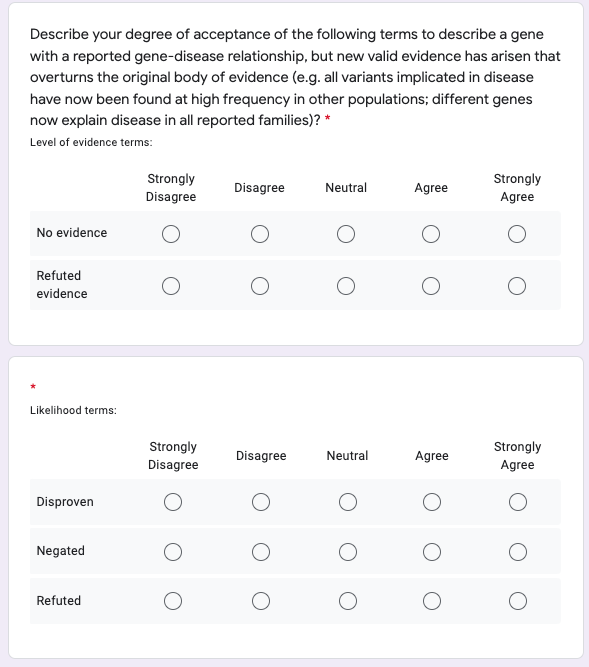

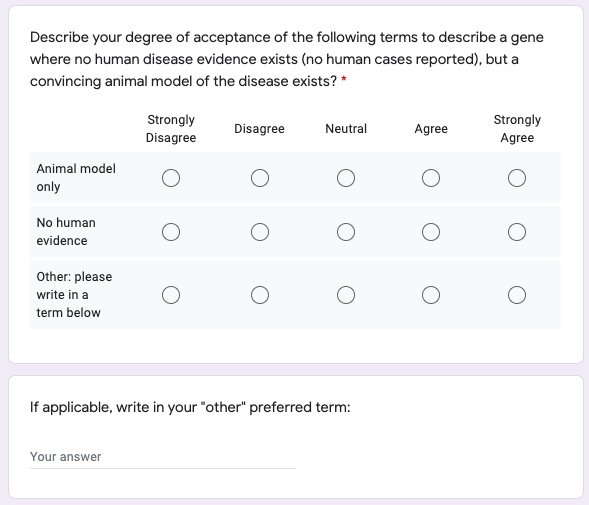

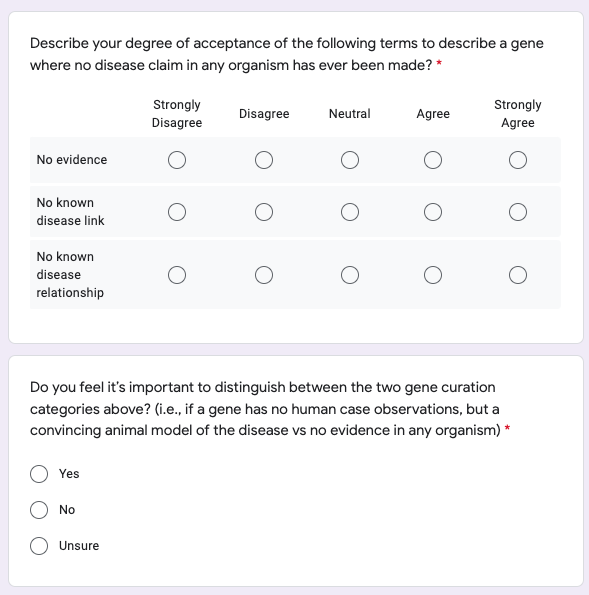

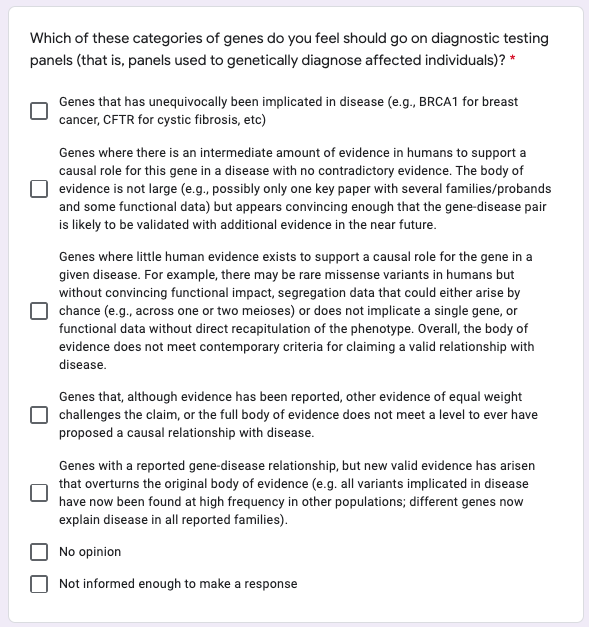

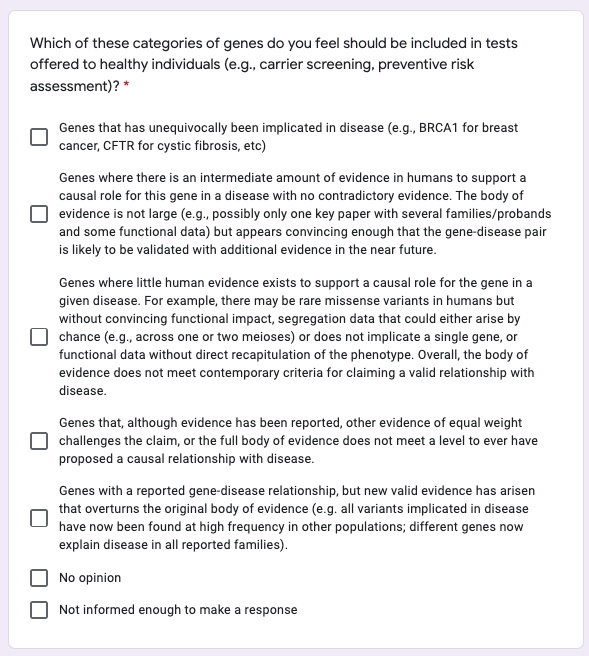

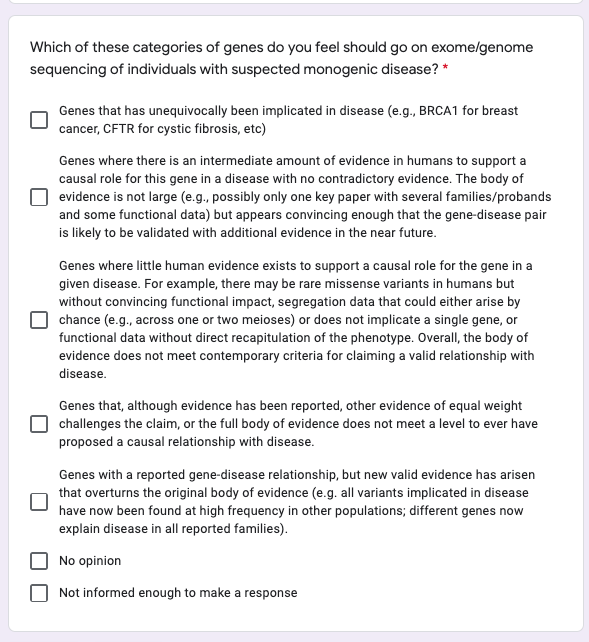

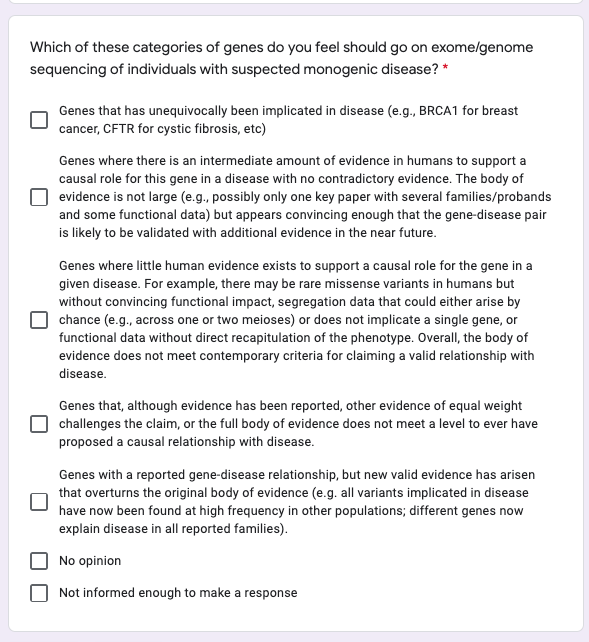

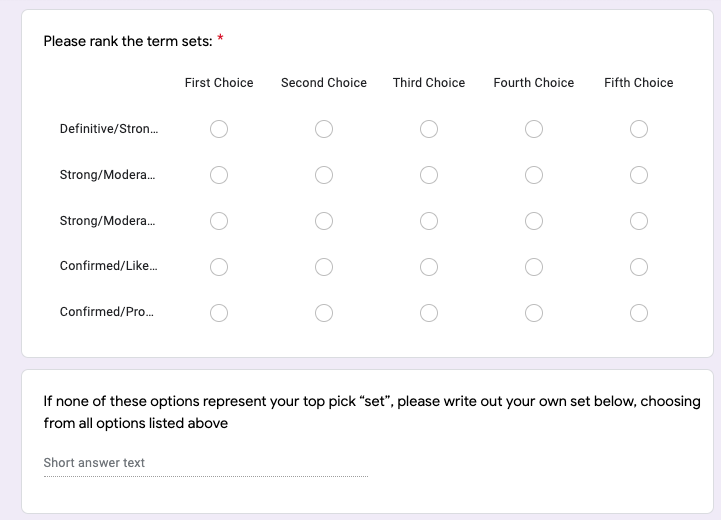

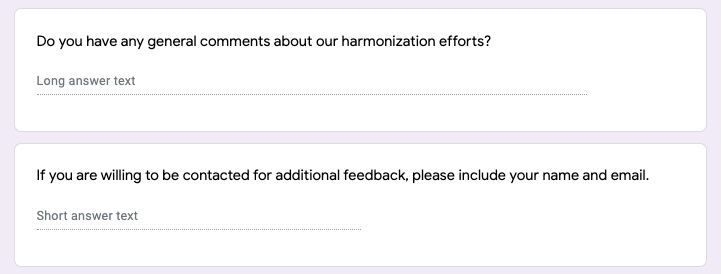
